# Genetic sensitivity analysis: estimating genetic confounding and environmentally mediated genetic effects using multiple exposures

**DOI:** 10.64898/2026.07.16.26358236

**Authors:** Leonard Frach, Frühling Rijsdijk, Laurie J. Hannigan, Frank Dudbridge, Jean-Baptiste Pingault

## Abstract

Polygenic scores are imperfect measures of the additive genetic effects of common genetic variants. The resulting measurement error biases estimates of quantities of interest in epidemiological analyses integrating polygenic scores. For example, how much of an exposure-outcome association is genetically confounded can be substantially underestimated when using polygenic scores alone. Here we present extensions to *Gsens*, a genetic sensitivity analysis, which aims to correct for such measurement error using both polygenic scores and heritability estimates. *Gsens* now allows for multiple exposures and estimates several quantities of interest, i.e. genetic confounding, adjusted residual association (net of genetic confounding), genetic overlap and environmentally mediated genetic effects. We present derivations and simulations showing how *Gsens* accounts for measurement error in the polygenic score; we also show how estimation may be affected by misspecifications of the causal structure between exposures. Applying *Gsens* in the Norwegian Mother, Father and Child Cohort Study (MoBa), we uncover, among other results, substantial genetic confounding in the associations between multiple known risk factors for attention deficit hyperactivity disorder (ADHD), such as low birth weight and temperament, and measures of ADHD in childhood. The updated *Gsens* R package offers multiple options, including for missing data handling and customisable syntax. Our extended version of *Gsens* is applicable to a broad range of substantive questions in multiple disciplines.

## Introduction

Since their emergence (1) polygenic scores (PGS) have been widely used in genetic and epidemiological studies. PGS are summary measures of the genetic propensity for a given phenotype, summarising the effects of thousands or millions of single nucleotide polymorphisms (SNPs) (2,3). Researchers have used PGS in several ways, including estimating genetic associations (e.g., associations between a PGS and an outcome), adjusting exposure-outcome associations for genetic confounding, and testing environmentally mediated genetic effects of PGS via mediators.

Integrating polygenic scores in epidemiological studies is not without challenges (4,5). Many of those challenges stem from the fact that PGS typically only capture a small fraction of the *heritability* of their corresponding phenotype. As a result, the use of PGS will lead to both under-and overestimation of quantities of interest (4). For example, several studies use PGS as covariates with the intent of adjusting exposure-outcome associations for genetic confounding (e.g., (6,7)). However, such studies will underestimate the magnitude of genetic confounding to the extent that the PGS does not capture the full heritability. Consequently, genetic confounding is not appropriately adjusted for, and the adjusted exposure-outcome association is overestimated.

The extent to which the predictive accuracy of a PGS for a phenotype (variance explained, denoted *R*^2^*_PGS_*) reflects the SNP heritability of that phenotype (variance explained by all common SNPs; *h*^2^*_SNP_*) can be described as reliability. We can define the reliability of the PGS as the ratio 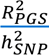, where a value of 1 describes a perfectly reliable PGS that matches the SNP heritability estimate (4). Conversely, measurement error can be defined as 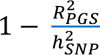. For example, in psychiatric genetics, reliability of PGS for, e.g., attention deficit hyperactivity disorder (ADHD) or schizophrenia ranges from 0.25 to 0.48 (4), whereas genetic effects on phenotypes such as height are now measured more reliably using PGS (8), e.g., with a reliability of 0.90 to 1 (for samples of European ancestry).

Initially, correcting for the imperfect reliability of PGS was implemented for bivariate PGS-trait associations (9,10). The first iteration of our approach, *Gsens* (Genetic sensitivity analysis) (11), focused on accounting for genetic confounding in the association between one exposure and one outcome. To that end, it used structural equation models (SEM) to integrate PGS and heritability estimates into a latent genetic factor. *Gsens* has since been widely used (12–19). For example, when adjusting for the imperfect reliability of PGS, genetic confounding entirely explained some associations between childhood maltreatment and internalising and externalising problems, whereas simply adjusting for PGS would have resulted in opposite conclusions (13). Importantly, this first iteration of *Gsens* only allowed us to consider one exposure at a time, while having multiples exposures is the most common scenario in epidemiological studies. For example, many parental exposures (e.g., parental education, maternal smoking during pregnancy and maternal pre-pregnancy body mass index (BMI)) have been associated with child ADHD in observational studies (20–22) but there is also evidence for genetic confounding between all those exposures and ADHD (11,23–25).

To address this limitation, we extend *Gsens* to simultaneously estimate multiple (continuous) exposure-outcome associations in the presence of genetic confounding. In addition, *Gsens* now enables the estimation of other quantities, including environmentally mediated genetic effects. For example, we can estimate how much of the genetic liability for ADHD is mediated by low birth weight. Gsens is customisable, supporting raw data or covariance-matrix inputs, multiple approaches to handling missing data, and a range of estimators and standard error options, including bootstrapping. We use derivations and simulations to show that *Gsens* recovers the true exposure-outcome associations under genetic confounding. We also consider how estimation is affected by misspecifications of the causal structure between exposures. Finally, using data from the Norwegian Mother, Father and Child Cohort Study (MoBa) (26,27), we apply *Gsens* to test the adjusted effects of birth weight and temperament (‘emotionality’ and ‘activity’) on later ADHD scores. Higher levels on both temperament dimensions and lower birth weight have been reported as predisposing factors for ADHD (28–31). As lower birth weight has also been associated with both temperament dimensions (32), we further assess whether effects of birth weight (and of genetic factors) on ADHD are mediated via temperament. We also apply *Gsens* to test the adjusted effects of parental education (both maternal and paternal), maternal smoking during pregnancy and maternal pre-pregnancy BMI on child ADHD scores.

## Results

### Method overview

We initially proposed a model using a PGS corrected for measurement error based on a heritability constraint (11), which can be set to a desired value (e.g., corresponding to the SNP heritability estimate of the trait which the PGS should index). For the ‘true’ underlying model, the constraint

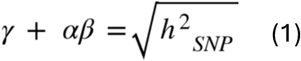

is derived from the direct effects of the genetic factor *G* on outcome *Y* (*_γ_*) and the indirect effects via exposure *X* (*α*β*) (**Figure 1a**).

**Figure 1.**
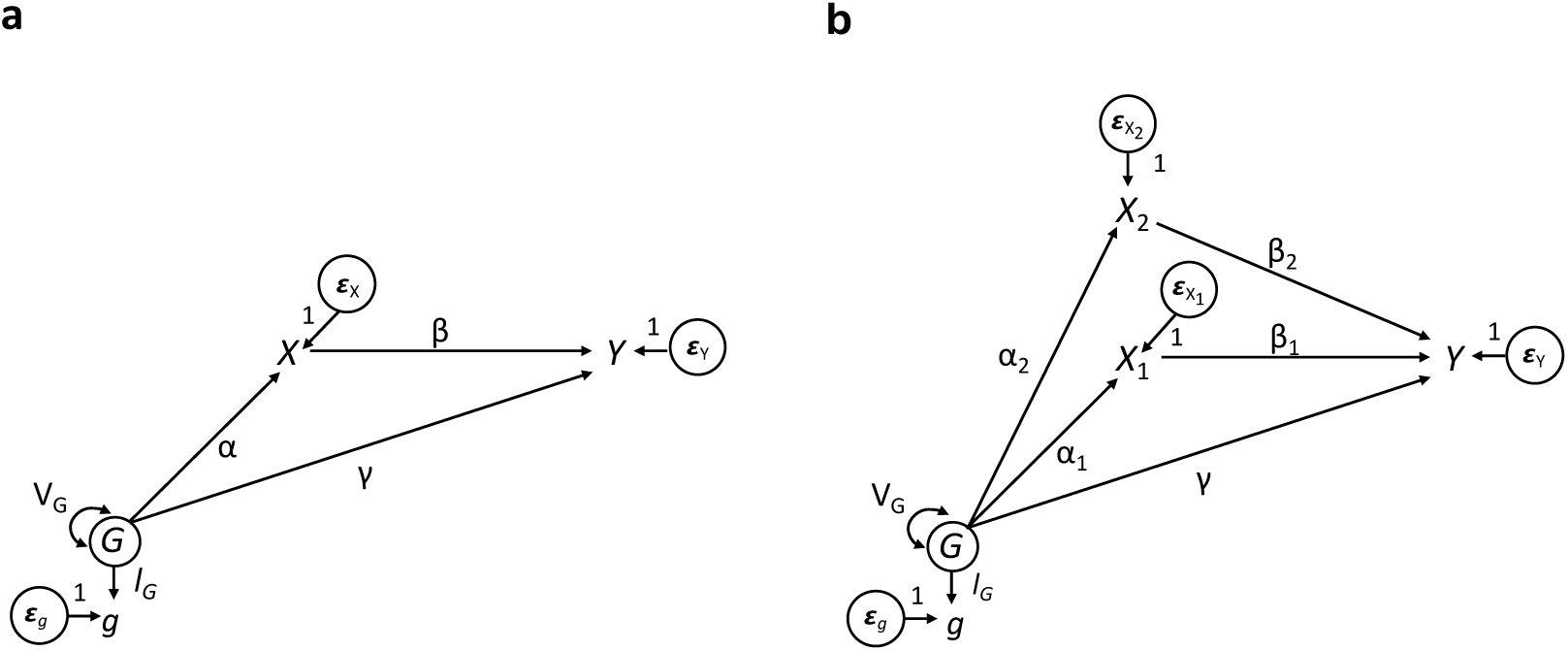
One exposure and multiple exposures models. *Note.* Figure 1 follows structural equation modelling conventions, including the use of circles to represent latent variables.

Here, we extend this initial work in three ways. First, we generalise the one exposure model to multiple exposures (example with two exposures in **Figure 1b**), which we identify through a generalisation of the heritability constraint. Second, based on this model with multiple exposures, we provide additional quantities of interest, including genetic overlap and environmentally mediated genetic effects. Third, we provide updated code that allows considerably more options in terms of input data, missing data approaches and robust estimators, among others.

Quantities of interest are formally defined in the *Methods* section. In brief, genetic confounding is the confounding effect resulting from the effect of the latent genetic factor on the exposure and the outcome (via paths *α* and *_γ_* in **Figure 1a**). For multiple exposures (**Figure 1b**), there are multiple genetic confounding paths for each exposure-outcome association (see *Methods* and Supplementary materials for derivations). The adjusted association is the exposure-outcome association remaining after removing genetic confounding (path *β* in **Figure 1a**). The environmentally mediated genetic effect is the effect of latent genetic factor on the outcome via the exposures, which can be captured jointly for all exposures or for each exposure separately (e.g., total mediated effect is *α*_1_*β*_1_ + *α*_2_*β*_2_ in **Figure 1b**). Genetic overlap for a given *X_i_*-*Y* association includes the genetic confounding terms for this *X_i_*-*Y* association but also the part of the effect of the exposure on the outcome that is genetic in origin (i.e. path *β* weighted by the variance in *X* explained by *G* in **Figure 1a**). Lastly, the ‘environmental association’ is the complement, i.e. the total association minus the genetic overlap.

### Simulation scenarios

We quantified bias, statistical power, and type 1 error rates of *Gsens* under different simulation scenarios with 1,000 iterations per scenario. For this, we simulated a PGS (*g*) for a trait (outcome) as a measure of the true genetic factor *G*, with different degrees of measurement error. We further varied the effect size of the exposure-outcome associations, residual correlations between exposures and heritability parameters (**Table 3**). As we present formal derivations in the two-exposure case (see methods), we included an additional exposure in power and bias simulations to showcase the generalisability of *Gsens* to multiple exposures. Bias, power and type 1 error were quantified for the main quantities of interest i) the adjusted exposure-outcome associations, ii) the environmentally mediated effect of *G* and for iii) genetic confounding. All simulation results can be found on [GitHub](https://github.com/LeonardFrach/Gsens/SimResults).

### Degree of measurement error impacts type 1 error rates, especially for single mediation effects

We simulated different scenarios with two exposures (see *Methods*) to calculate type 1 error rates for different quantities of interest, including the adjusted exposure-outcome association, mediation and genetic confounding estimates. For the adjusted association, type 1 error was examined by setting one parameter to zero, i.e., one of the simulated adjusted associations between the two exposures and the outcome. For mediation, type 1 error for a given mediation effect, e.g. *α*_1_*β*_1_, was examined if either the true *α* or the true *β* path was simulated to be zero. As the genetic confounding includes multiple terms (see equation 4), type 1 error for genetic confounding estimates could be examined if any parameter of the included term was simulated to be zero (i.e., for *gc*_1_ this was the case if *α*_1_ was 0, since *_γ_* was never 0). In general, type 1 error rates for all types of estimates (adjusted exposure-outcome associations, mediation effects and genetic confounding) were ≤ 0.05 across most simulation scenarios. Type 1 error rates were generally very close to 0.05 for adjusted associations, with estimates within range of random error over 1,000 simulations (95% CI = [0.036, 0.064]) across almost all parameter combinations (**Figure 2a**). Similarly, the type 1 error rate for the genetic confounding estimates was close to 0.05 for most scenarios (**Figure 2b**). Type 1 error rates were slightly inflated when there were null effects of *G* on exposures (i.e., no true confounding) but exposures were correlated (error rates of up to 0.066). For the total mediation effect (that is, when the true simulated effects of the genetic factor on all exposures were zero (*α_i_* = 0) *or* when the true effects of the exposures on the outcome were zero (*β_i_* = 0)), we found type 1 error rates ≤ 0.05 across most simulation scenarios (**Figure 2c**). For individual mediation paths (*m*_1_ *or m*_2_), type 1 error rates were somewhat inflated and increased to up to 0.085 for some scenarios with large measurement error ([inine]), especially when heritability was higher (see **Figure 2d**). This was observed in cases where, for example, one mediation path was simulated to be 0 (if *α*_1_ or *β*_1_ were 0) but paths involving other exposures were positive (e.g. *α*_2_ or *β*_2_).

**Figure 2.**
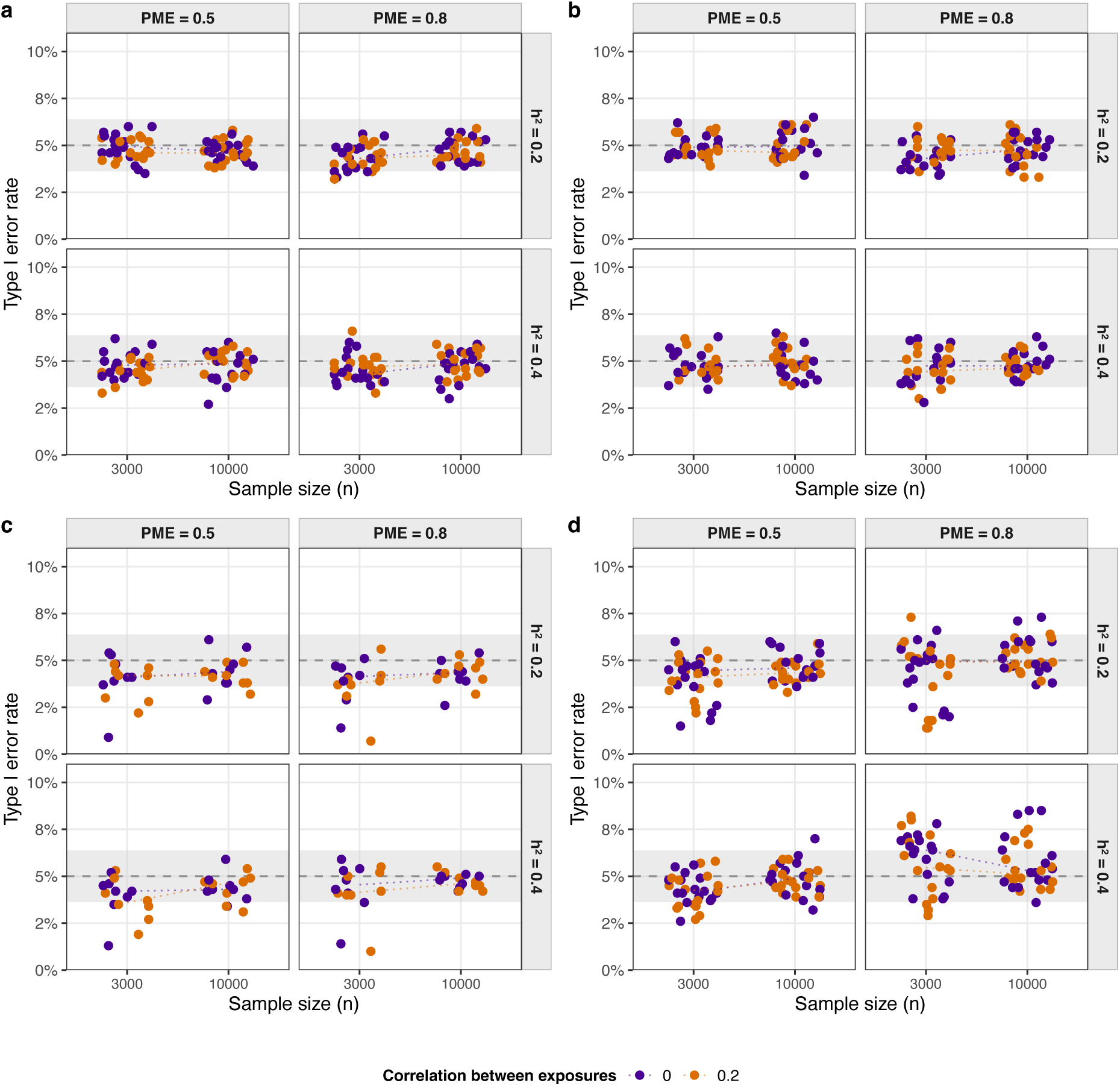
Type 1 error rates for adjusted associations, genetic confounding and mediation. *Note.* Type 1 error rates for scenarios with two exposures where **a)** the true adjusted associations (*β*_1_ or *β*are 0, **b)** the true genetic confounding (*gc*_1_ or *gc*_2_) is 0, **c)** the true total mediation is 0 and where **d)** the true mediation pathways (*m*_1_ or *m*_2_) are 0. Only one parameter at a time was set to 0, whereas the others were non-null (see *Methods* for overview).

### Bias in genetic confounding depends on effect sizes of exposure-outcome associations

Bias was estimated as the discrepancy between the simulated effect and the estimated effect. Bias for estimates of the adjusted associations between exposures and the outcome was very low (|bias| < 0.005) in all scenarios. For mediation effects, bias estimates were also very small (|bias| ≤ 0.001), which is expected as we did not model any unobserved nongenetic confounding in the underlying theoretical model. Note that the estimated bias did not depend on the residual correlation between exposures, which is explicitly modelled in the *Gsens* method. For genetic confounding, we observed somewhat larger bias if the true exposure-outcome association was very small (e.g., *β_i_* = 0.05). Notably, bias in genetic confounding for an exposure (e.g. *X*_2_-*Y*) was elevated if parameters involving another exposure (e.g. *α*_1_ and *β*_1_) were larger. This is because genetic confounding for *X*_2_ also includes those parameters as can be seen with the backdoor path *α*_2_*α*_1_*β*_1_ in **Figure 1b**.

### Limited power to detect mediation effects for small sample size when measurement error is large

In general, *Gsens* was sufficiently powered to detect small and moderately sized exposure-outcome associations for the tested sample sizes under different degrees of genetic confounding (**Figure 3a**). Occasionally, power was below 0.80 if sample size was small (*N* = 3,000) and measurement error was large 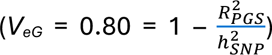. Similarly, power to detect significant genetic confounding was below 0.80 for scenarios with smaller sample size (*N* = 3,000) and larger measurement error, especially if the true associations between *G* and exposures were very small (e.g., *α* = 0.10) (**Figure 3b**). Overall, for the total mediation, statistical power was well above 0.80 across all scenarios (**Figure 3c**), even when some effects were very small (e.g., *β*_2_ = 0.05), as the total mediation also depends on the larger mediation effects via other exposures. In contrast, power to detect single mediation pathways was again limited for scenarios with smaller sample size and larger degrees of measurement error (**Figure 3d**). Overall, as expected, small sample size and larger degrees of measurement error in *g* negatively affected statistical power. The size of heritability estimates did not seem to largely affect power in our simulations, though in reality measurement error and heritability estimates are likely correlated, which can influence statistical power (33).

**Figure 3.**
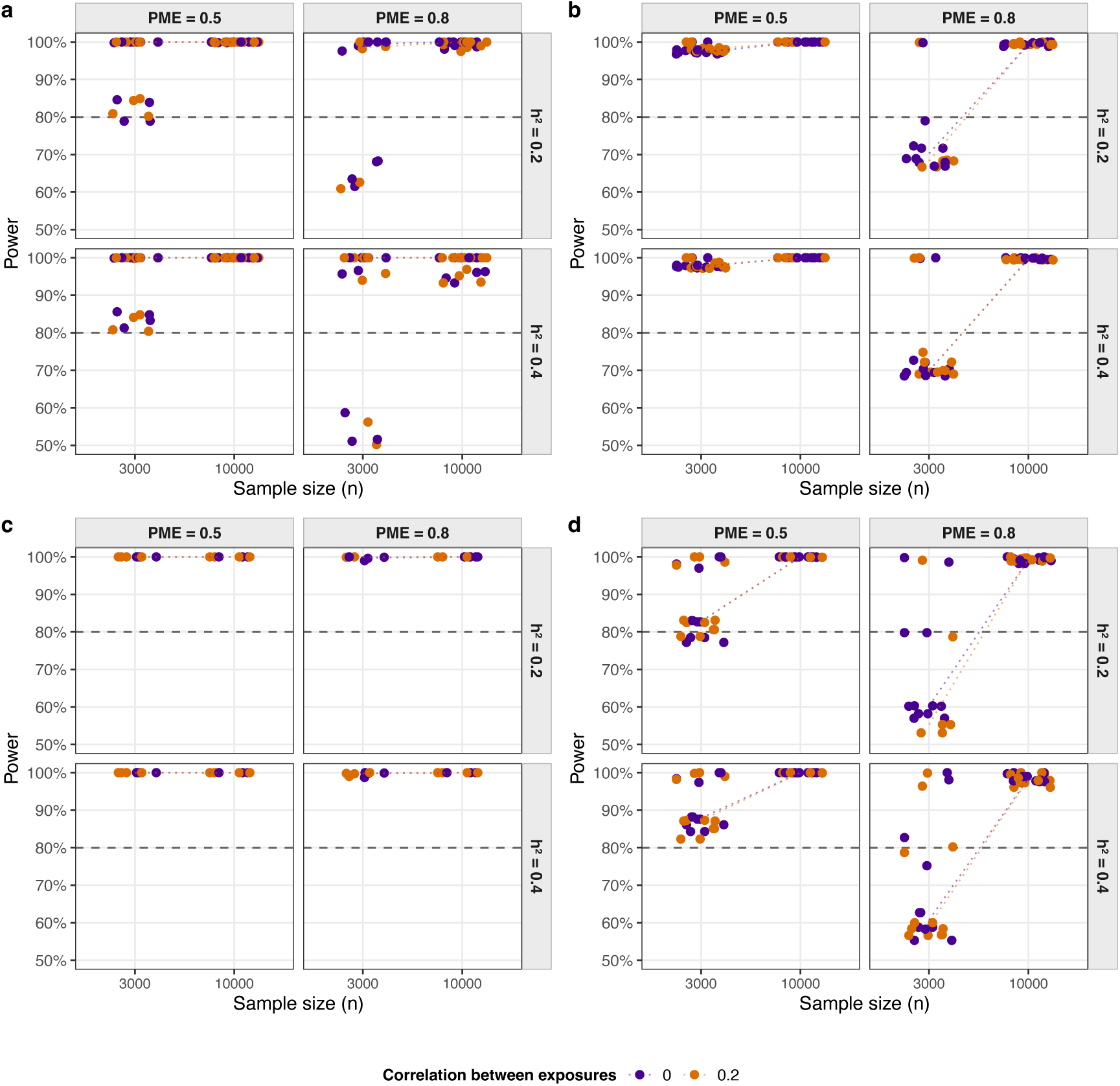
Statistical power for adjusted associations, genetic confounding and mediation. *Note.* Power for detecting non-null effects for **a)** adjusted exposure-outcome associations, **b)** genetic confounding, **c)** total mediation and **d)** individual mediation pathways

### Model misspecifications

Although *Gsens* allows for residual covariances between exposures, it is agnostic to the nature of this residual covariance, e.g., due to a common cause of *X_i_* and *X_j_*, (**Figure 4a**) or due to a causal effect between *X_i_* and *X_j_*. To test the impact of model misspecifications, we simulated scenarios where a true causal effect between two exposures in the underlying model was not specified in the fitted *Gsens* model (**Figure 4b**). For this, we simulated three exposures and a causal effect 8 of exposure *X*_1_ on exposure *X*_2_ with a size of 0.25 (**Table 4**). As also shown in the derivations in the **Supplementary Information** (section 3), the simulations revealed that the total mediation effect was not biased (mean bias of close to 0) and both *m*_1_ and *m*_3_ had a bias of 0. However, as expected from the derivations, *m*_2_ showed a small bias of 0.005, as the path *a*_2_ was overestimated by disregarding mediator *X*_1_. Technically, the mediation path of *X*_1_ via *X*_2_ (8) is still a true mediation effect via *X*_2_, but not a ‘direct’ mediation effect of *G* on *Y*.

**Figure 4.**
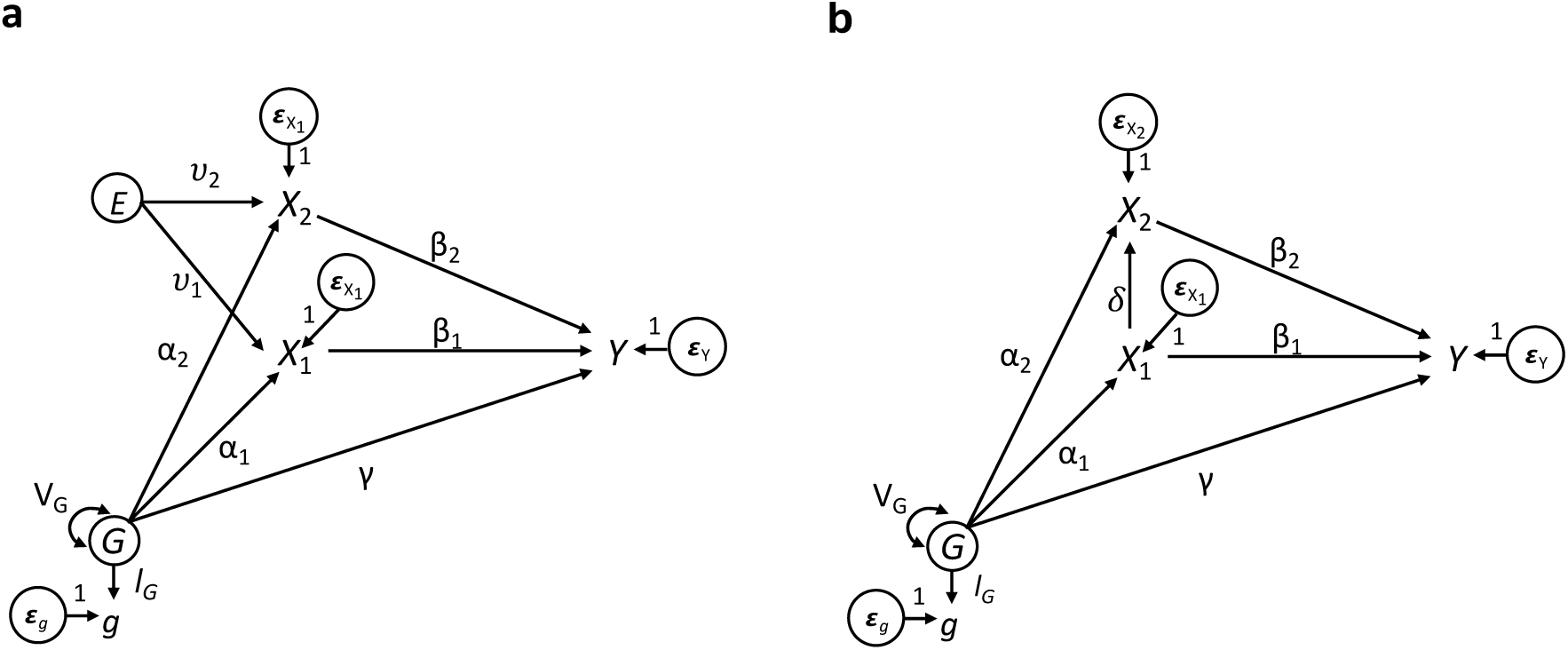
Potential misspecifications of associations between exposures. *Note.* **a)** Unobserved environmental confounder (*E*) with causal effects on both exposures, resulting in a residual covariance between exposures. **b)** Causal effect of exposure 1 on exposure 2, which will also result in a residual covariance between exposures if this causal path is not modelled.

### Empirical applications

We applied the extended *Gsens* approach to data from the Norwegian Mother, Father and Child Cohort Study (MoBa) (26,27). We tested the following two examples adjusting for genetic confounding (**Figure 6**): i) joint effects of birth weight and the temperament dimensions ‘emotionality’ and ‘activity’ at age 3 years on ADHD scores at age 8 years and ii) the joint effects of parental education, maternal smoking during pregnancy and maternal BMI during pregnancy on offspring ADHD scores at age 8 years. See the *Methods* section for details on the sample and measures.

For comparison, we initially fitted linear models for both application examples to assess the phenotypic associations of the exposures with ADHD in the genotyped subsample with complete data, that is, *N* = 26,307 for example one and *N* = 21,730 for example two. As expected, phenotypic associations between all exposures and ADHD were significant (**Supplementary Table S2**).

Running *Gsens* (using the gsensY() function) to adjust associations between birth weight, temperament and ADHD for genetic confounding revealed moderate and significant genetic confounding between all exposures and ADHD (see **Table 1**). Despite genetic confounding, small to moderate adjusted associations between all exposures and ADHD remained significant, albeit with some marked attenuation. For reference, for birth weight, genetic confounding explained about one third of the total observed association between birth weight and ADHD, whereas for the temperament dimension of activity, genetic confounding explained more than half of the total observed association. In general, genetic overlap estimates were highly similar to genetic confounding estimates, which is to be expected unless there are very strong adjusted exposure-outcome associations. Furthermore, there was a small significant mediation of genetic effects on ADHD via all exposures (**Table 1**). Results using the restricted sample of unrelated individuals were fairly consistent (*N* = 17,185; see **Supplementary Table S3**). Specifying a causal effect of birth weight on the two temperament dimensions revealed a significant negative effect of birth weight on emotionality (*β* = –0.024, 95% CI = [–0.037, –0.012]) and no effect on activity (*β* = –0.006, 95% CI = [–0.021, 0.009]). The serial mediation effect birth weight → emotionality → ADHD was also significant (**Supplementary Tables S4-5**). As expected, associations between the latent genetic factor and sex were close to zero and non-significant, which shows that nongenetic covariates can be integrated in the model as they are not influential for the heritability equation. Alternatively, the effects of nongenetic covariates can also be regressed out prior to using *Gsens*.

**Table 1.** Application 1. Outcome ADHD, exposures: birth weight, emotionality and activity.

|  | *Std Est* | *SE* | *CI lower* | *CI upper* | *Z* | *p-value* |
| --- | --- | --- | --- | --- | --- | --- |
| Sex | -0.190 | 0.009 | -0.208 | -0.172 | -20.44 | $7.6 \times 10^{-93}$ |
| Adjusted association:<br>Birth weight | -0.033 | 0.009 | -0.050 | -0.015 | -3.63 | $2.8 \times 10^{-04}$ |
| Adjusted association:<br>EAS Emotionality | 0.170 | 0.009 | 0.153 | 0.187 | 19.42 | $5.7 \times 10^{-84}$ |
| Adjusted association:<br>EAS Activity | 0.062 | 0.010 | 0.042 | 0.082 | 6.12 | $9.3 \times 10^{-10}$ |
| Mediation via BW | 0.001 | 0.000 | 0.001 | 0.002 | 3.10 | $1.9 \times 10^{-03}$ |
| Mediation via EAS_Emo | 0.009 | 0.003 | 0.003 | 0.015 | 2.83 | $4.7 \times 10^{-03}$ |
| Mediation via EAS_Act | 0.014 | 0.002 | 0.011 | 0.017 | 9.22 | $2.9 \times 10^{-20}$ |
| Total mediation | 0.019 | 0.006 | 0.008 | 0.031 | 3.41 | $6.6 \times 10^{-04}$ |
| Genetic confounding BW | -0.016 | 0.008 | -0.032 | -0.001 | -2.12 | 0.034 |
| Genetic confounding EAS_Emo | 0.019 | 0.007 | 0.005 | 0.033 | 2.62 | $8.8 \times 10^{-03}$ |
| Genetic confounding EAS_Act | 0.082 | 0.008 | 0.066 | 0.098 | 10.02 | $1.2 \times 10^{-23}$ |
| Genetic overlap BW | -0.017 | 0.008 | -0.032 | -0.001 | -2.12 | 0.034 |
| Genetic overlap EAS_Emo | 0.020 | 0.008 | 0.005 | 0.035 | 2.56 | 0.010 |
| Genetic overlap EAS_Act | 0.085 | 0.008 | 0.069 | 0.101 | 10.15 | $3.5 \times 10^{-24}$ |

*Note.* All associations between G and covariate sex were close to zero and not significant (as expected), hence they are omitted from the table. 95% confidence intervals are specified. BW = birth weight, EAS = Emotionality, Activity and Shyness Scale (measuring temperament dimensions)

For the second application example of *Gsens*, we regressed out the effects of the covariates (child sex and parental age) on all exposures. This was done because both maternal and paternal age were significantly negatively correlated with the child PGS for ADHD (*r* = –0.056, 95% CI = [–0.063, –0.049] and *r* = –0.032, 95% CI = [–0.039, –0.025] for maternal and paternal age, respectively) and therefore would have influenced the heritability equation if added as additional exposures (covariates).

We found significant genetic confounding effects between all parental exposures (maternal and paternal education, maternal smoking during pregnancy and maternal pre-pregnancy BMI) and ADHD (**Table 2**). For the association between maternal and paternal education and ADHD, genetic confounding estimates were larger than the total observed phenotypic associations. Thus, after adjusting for the negative genetic confounding, associations between parental education and ADHD flipped sign and were positive, and although small, significant. The small positive association between maternal smoking at the start of the pregnancy remained significant, but the associations between maternal smoking after the pregnancy was known and maternal pre-pregnancy BMI and ADHD were not significant after adjusting for genetic confounding (**Table 2**). Restricting the analytic sample to unrelated individuals confirmed the genetic confounding between all exposure-ADHD associations, albeit with expectedly lower statistical power (*N* = 14,156; **Supplementary Table S6**).

**Table 2.** Application 2. Outcome ADHD, exposures: parental education, maternal smoking during pregnancy and pre-pregnancy BMI.

|  | *Sdt Est* | *SE* | *CI lower* | *CI upper* | *Z* | *p-value* |
| --- | --- | --- | --- | --- | --- | --- |
| Maternal education | 0.034 | 0.013 | 0.008 | 0.060 | 2.57 | 0.010 |
| Paternal education | 0.040 | 0.014 | 0.013 | 0.068 | 2.88 | 4.0 x 10<sup>-03</sup> |
| Smoking during pregnancy (start) | 0.031 | 0.014 | 0.003 | 0.058 | 2.21 | 0.027 |
| Smoking during pregnancy | 0.002 | 0.014 | -0.025 | 0.030 | 0.18 | 0.858 |
| Maternal BMI (pre-pregnancy) | -0.012 | 0.011 | -0.034 | 0.010 | -1.07 | 0.284 |
| Genetic confounding Mat. edu | -0.112 | 0.012 | -0.134 | -0.089 | -9.63 | 6.1 x 10<sup>-22</sup> |
| Genetic confounding Pat. edu | -0.123 | 0.012 | -0.147 | -0.098 | -9.92 | 3.4 x 10<sup>-23</sup> |
| Genetic confounding SDP (start) | 0.052 | 0.009 | 0.035 | 0.069 | 6.02 | 1.8 x 10<sup>-09</sup> |
| Genetic confounding SDP | 0.050 | 0.009 | 0.033 | 0.068 | 5.73 | 9.9 x 10<sup>-09</sup> |
| Genetic confounding Mat. BMI | 0.060 | 0.009 | 0.042 | 0.079 | 6.54 | 6.0 x 10<sup>-11</sup> |

*Note.* This table omits the genetic overlap estimates, which are highly similar to the respective genetic confounding estimates. The first four rows show the adjusted associations between exposures and ADHD after adjusting for genetic confounding. edu = education, mat. = maternal, pat. = paternal

## Discussion

We have developed a flexible method, *Gsens*, which integrates polygenic scores and heritability estimates to quantify genetic confounding and genetic overlap, adjust exposure-outcome associations for genetic confounding, and estimate environmentally mediated genetic effects. This method, which allows for multiple exposures, has a broad scope of applications across disciplines and can be used to remedy some concerning biases that can arise from the use of polygenic scores in epidemiological models (4). Using *Gsens*, we demonstrated, for example, that the associations between known precursors of ADHD and ADHD scores in childhood are genetically confounded.

### Polygenic scores in epidemiological studies

A growing body of studies are using polygenic scores in epidemiological models to answer a variety of questions. A first set of studies have used polygenic scores to adjust exposure-outcome associations, in an attempt to account for genetic confounding (6,7). A second set of studies have explicitly tested mediation of PGS effects on outcomes via exposures of interest (34,35). For instance, the association between a PGS for ADHD and general psychopathology has been shown to be partially mediated by cognitive abilities (36). However, in both set of studies, simply modelling PGS can lead to substantial biases, for example by underestimating the extent of genetic confounding and thus overestimate the adjusted exposure-outcome association net of confounding; or by overestimating the role of a mediator in explaining genetic effects on the outcome (see reference (4) and **Supplementary Information**). *Gsens* offers a flexible tool to address those biases. *Gsens* now also offers the possibility of using the rich set of options typically available within SEM, including a range of estimators (e.g. maximum likelihood), missing data options (e.g. multiple imputation), multiple options for standard errors (e.g. robust or bootstrapping). It also allows for the flexible customisation of model specification, e.g., adding additional causal effects between exposures (e.g., see **Supplementary Tables S4-5**) or modelling measurement error in the exposures and outcomes by modelling corresponding latent factors. While *Gsens* cannot adjust for all bias, for example bias arising from unobserved nongenetic confounding, it should reduce bias in the exposure-outcome associations compared to simply using PGS or not adjusting for genetic confounding at all.

### Simulation results

Our results showed that *Gsens* is sufficiently powered to detect small adjusted exposure-outcome associations, mediation effects and genetic confounding in sample sizes of a few thousand participants. Genotyped datasets of this size are now commonly available and widely accessible. Furthermore, our simulation results showed that type 1 error rates are controlled (< 0.05) in most scenarios but that error rates can be larger when estimating mediation effects (total mediation and individual pathways) if the degree of measurement error is larger (0.80) and the sample size is comparably small (*N* = 3,000). The simulations on model misspecifications (i.e., omitting a causal effect between exposures in the model specification) confirmed the derivations in the **Supplementary Information** section 3, showing that exposure-outcome associations, the total mediation effect and thus the genetic confounding estimates are unaffected.

### Predisposing factors for ADHD are not fully explained by genetic confounding

Using *Gsens* in our empirical examples, we detected significant genetic confounding between all predisposing factors and ADHD, in addition to some significant adjusted exposure-outcome associations. The adjusted association between birth weight and ADHD is consistent with findings from genetically informed studies, e.g., sibling comparisons or twin designs (37,38). Genetic confounding in the associations between the two temperament dimensions and ADHD is in line with findings from a longitudinal twin study, which showed that the associations between temperament and ADHD are genetically influenced (39). While adjusted associations between both temperament dimensions and ADHD remained significant, unobserved environmental confounding or shared method bias could still explain these adjusted associations between mother-reported temperament and mother-reported ADHD (39,40). Interestingly, we found a significant mediation effect of birth weight on ADHD via emotionality independent of genetic confounding, which provides some insights into direction of effects of early predisposing factors for ADHD.

We found significant genetic confounding for all parental exposure-ADHD associations (**Table 2**). After adjusting for genetic confounding, the associations with maternal pre-pregnancy BMI and smoking during pregnancy (after the pregnancy was known) were not significant. This suggests that associations of maternal pre-pregnancy BMI and smoking during pregnancy with ADHD may not be causal but mainly explained by genetic confounding. This is not surprising, as both BMI and smoking behaviour share genetic effects with ADHD (41). Although we found a nominal significant adjusted association with maternal smoking at the start of the pregnancy, this association was not robust in sensitivity analyses, in line with converging evidence from multiple causal inference methods showing that the effect of smoking during pregnancy on ADHD is not likely to be causal (42).

Unexpectedly, we found that compared to the phenotypic associations between parental education and offspring ADHD, the adjusted parental education-ADHD adjusted associations in *Gsens* were significant but in the opposite direction (positive). This could mean that, after adjusting for genetic effects (genetic confounding estimates were negative and comparably large), mothers and fathers who are more highly educated have offspring with higher levels of ADHD. This could result from a combination of both rater biases (highly educated mothers may be more likely to detect and report problematic behaviour of their offspring) and selection bias, as genotyped MoBa families tend to have higher education than the whole MoBa sample and the general Norwegian population. Furthermore, in the subsample of unrelated individuals, only the association between paternal education and ADHD remained significant, with a 50% attenuation of the association between maternal education and ADHD (**Supplementary Table S6**). Similar unexpected findings have been reported of positive familial genetic effects of maternal PGS for education and child externalising problems such as conduct problems (43).

We note that, when examining the associations between parental factors and offspring ADHD, we only included the child PGS (or genetic factor) for ADHD in the model. We thus only account for transmitted genetic effects, which is sufficient to account for narrow sense genetic confounding. However, potential confounding effects of parental non-transmitted alleles is not accounted for in this model. From the offspring perspective, the confounding by non-transmitted genetic effects can be considered as ‘environmental effects’ on the offspring and thus part of the unobserved environmental confounding that is not accounted for in *Gsens*.

### Strengths of Gsens

Measurement error of PGS is now more widely discussed and different approaches have emerged to adjust genetic associations involving PGS (4,44). To date, these approaches have not specifically addressed genetic confounding or mediation analysis (9,10). However, recent methods have been proposed to account for genetic confounding using genome-wide data, as opposed to using PGS as summary measures (45), e.g. using mediation framework (46) or to estimate mediation of genome-wide effects on an outcome via a single exposure (47). In contrast to *Gsens*, these methods currently allow assessment of just one exposure-outcome association at a time. Notably many researchers in applied fields (e.g., psychology, social sciences) may not be familiar with preprocessing and analysing genome-wide data, whereas many cohort studies such as MoBa, the Avon Longitudinal Study of Parents and Children (48) or the British Cohort studies now provide pre-computed PGS, which makes the application of *Gsens* straightforward for researchers without advanced training and background in genetics.

### Limitations

This study has some limitations. First, as in all simulation studies, we did not test all (theoretically) possible parameter combinations of heritability, effect size, sample size and measurement error. With respect to computational efficiency and interpretability of results, we focused on sample sizes that are frequently used in studies involving PGS and on effect sizes that are typically observed or expected when modelling complex traits (that is, no ‘large’ effect sizes of single exposures). Similarly, this applies to parameters for heritability and measurement error. However, we did not model exposures or outcomes that are only influenced by a few causal genetic variants (as opposed to a polygenic architecture), as most past and future applications of *Gsens* are to be expected in studies of complex traits. For more biological exposures, where a few causal variants with strong effects can be identified (e.g. metabolites), other methods such as Mendelian randomisation may be preferred.

## Conclusions and future directions

*Gsens* is a flexible method to test multiple exposure-outcome associations, while correcting for measurement error resulting from the use of PGS. *Gsens* should be seen as complementary to other methods and triangulation with other designs is necessary to draw conclusions about causality. Given the high customisability, future extensions of *Gsens* can be developed, e.g., to include categorical exposures and outcomes (e.g., binary outcomes) or to test gene-environment interactions.

## Methods

### Definition of quantities

In a standardised model, we now present the formulae for the different quantities. We refer to quantities with respect to the theoretical model (**Figure 5a**) and provide the estimated *Gsens* model (**Figure 5b**) for further reference (see **Supplementary Information**). First, we define *heritability* using *k* exposures as

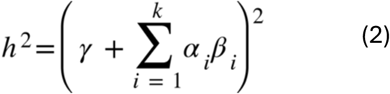

**Figure 5.**
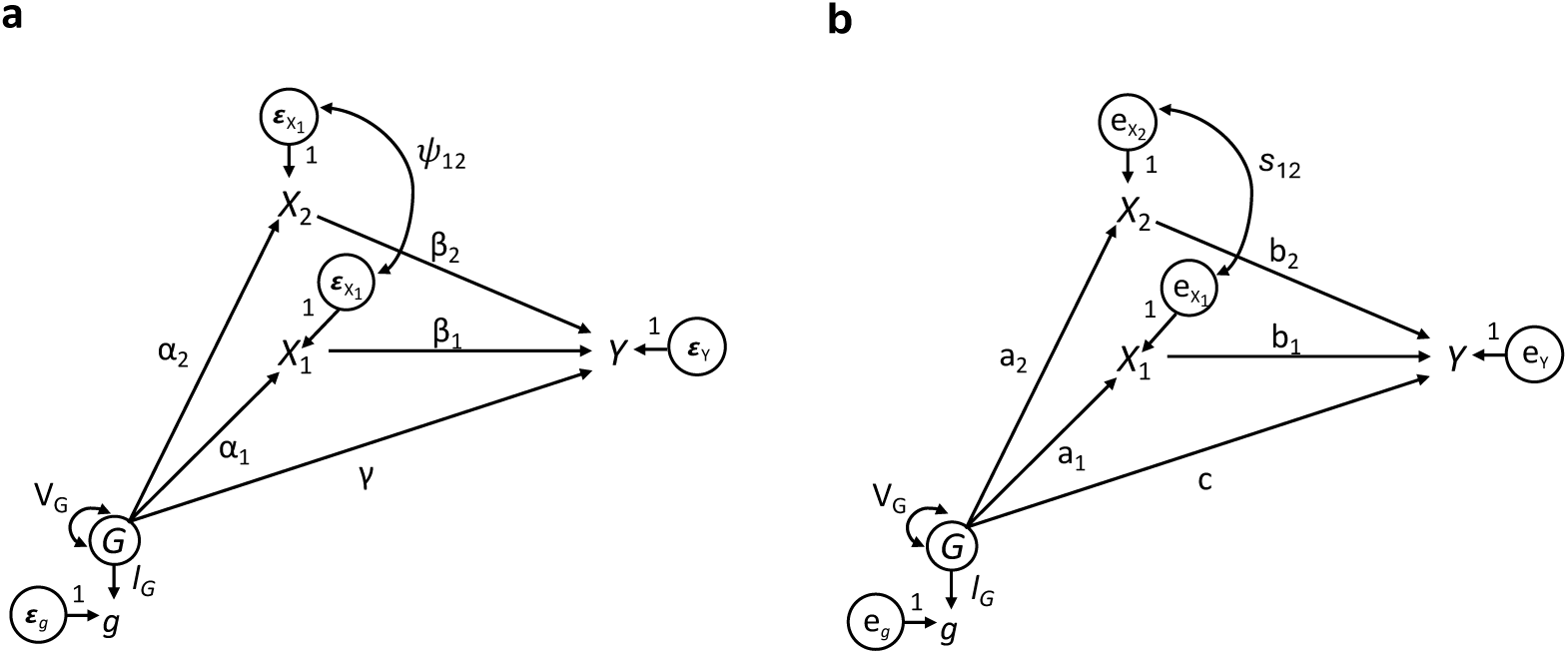
Theoretical and fitted Gsens model using a latent genetic factor. *Note.* **a)** Theoretical model with residual covariance between exposures (see section below and Figure 2 for scenarios that could create this residual covariance). *α* = direct effects of genetic factor *G* on exposures, *β* = direct effects of exposures on the outcome, *ψ* = direct effect of *G* on the outcome; *l_G_* = loading of PGS (*g*) on *G*, which corresponds to the square root of the reliability of *g*. **b)** Fitted *Gsens* model where *g* corresponds to the PGS, and the residual covariance between exposures is accounted for, irrespective of the underlying causal model.

where *ψ* is the direct effect of *G* on outcome *Y*, ***a*** is a vector of length *k* containing the effects of *G* on all exposures, and ***b*** is a vector of length *k* containing the effects of all exposures on outcome *Y* (see section 1 of the **Supplementary Information**). For two exposures, this is

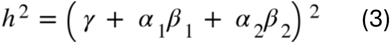

In addition, 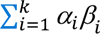 describes the total mediation effect of genetic effects on *Y* that are mediated via all exposures, and *α_i_β_i_* is the mediation via a single exposure *i*. Furthermore, we define the genetic confounding paths for *k* exposure-outcome association as

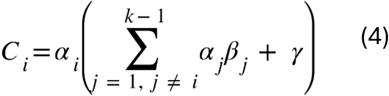

For the example with two exposures, genetic confounding for the *X*_1_-*Y* association will be

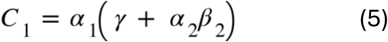

The genetic overlap for each exposure-outcome association contains the respective genetic confounding paths and part of the causal effect of exposure *i* that originates in *G*. This can be expressed as

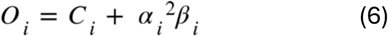

Finally, as shown in the **Supplementary Information**, the total association between an exposure *i* and *Y*, e.g., the correlation when using the standardised model, can be described as

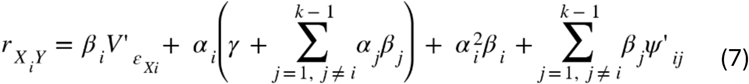

where *k* is the number of exposures and *V’_εXi_* is the standardised residual variance of exposure *i* and where *ψ.’_ij_* is the standardised residual covariance (correlation) between exposures *i* and *j*. As shown in the **Supplementary Information**, the terms that are scaled by *b_i_* can be summarised to the variance of *X_i_*, which is 1 when standardised and thus

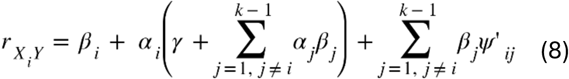

As mentioned, the residual covariance between exposures can be caused by an unobserved nongenetic confounder *E* (section 4 of the **Supplementary Information**) or by a causal effect between exposure *X_i_* and exposure *X_j_* (section 3). Importantly, the estimated *b_i_* is the adjusted association between *X_i_* and *Y* after adjusting for genetic confounding and the residual covariances with all other exposures. This may, however, not be interpreted as a causal effect, given potentially unobserved nongenetic confounding in the *X_i_-Y* association. We show in the application example that measured confounders or non-genetic covariates (e.g., sex) can be integrated in the model by either including them as additional exposures or by residualising the affected exposures (regressing out the effects of the covariates on the exposures).

#### Software implementation

Beyond the derivations of the presented quantities, we provide an updated *Gsens* R package [v0.1.6](https://github.com/LeonardFrach/Gsens), which estimates these additional quantities in both the single and the multiple exposure scenarios. The *Gsens* package was developed using R v4.3.1 (49) and is wrapped around the SEM *lavaan* package v0.6.20 (50).

#### Heritability constraint

In addition to the heritability constraint (see Method overview) other approaches use a ratio of

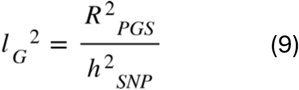

for model identification, where *l_G_* represents the loading of the PGS on the true genetic factor (9,44). We previously showed that this ratio constraint assumes no measurement error in outcome *Y* (4), and we show in the **Supplementary Information** (section 5) that under this assumption, and if *h*^2^*_Y_* = *h*^2^*_SNP_*, the heritability constraint and the loading constraint are equivalent. We also show that the equivalence of both constraints holds in the multiple exposure case (see **Supplementary Information** section 5). In addition to the main *Gsens* model using the heritability constraint, we thus also added an option to use a loading constraint instead.

Importantly, the initial *Gsens* R package and publication (11) included three functions: gsensX(), gsensY() and gsensXY(), adjusting for the genetic factors of *X*, *Y* and both, respectively. Here, we chose to extend the gsensY() function, given previous findings (4,11,51,52) that using a genetic factor for *X* can lead to bias amplification in the presence of colliders in the model (see also section 6 of the **Supplementary Information**). GsensY() models a genetic factor capturing all common (additive) genetic effects on the outcome *Y*, either directly or via any exposures in the model. Furthermore, this approach avoids the potential issue of overfitting when scaling to scenarios of dozens of exposures (which would require including dozens of genetic factors in the other two functions).

#### Multiple (correlated) exposures

Using the case of two (potentially correlated) exposures, we additionally explore some misspecifications. **Figure 4** shows two different underlying models, which both result in a residual correlation between exposures. This correlation can be caused by an unobserved common factor (**Figure 4a**) or by a causal effect of one exposure on the other (**Figure 4b**). We show that, if there is a causal relationship between the different exposures, the estimate of the total environmentally mediated genetic effects (i.e. jointly by all exposures) remains unbiased, but effects mediated by specific exposures may be biased unless explicitly specified (see section 3 of the **Supplementary Information**).

#### Simulations

We ran different sets of simulations testing power, type 1 error and bias of the *Gsens* method under different scenarios. For this, we simulated a PGS for the outcome, with different degrees of measurement error of the PGS (*g*) as a measure of the true genetic factor *G* and different effect sizes for exposure-outcome associations and heritability (**Table 3**). We provide derivations for the case of two exposures and thus included three exposures in simulations to test how the method can be generalised to more than two exposures.

**Table 3.**
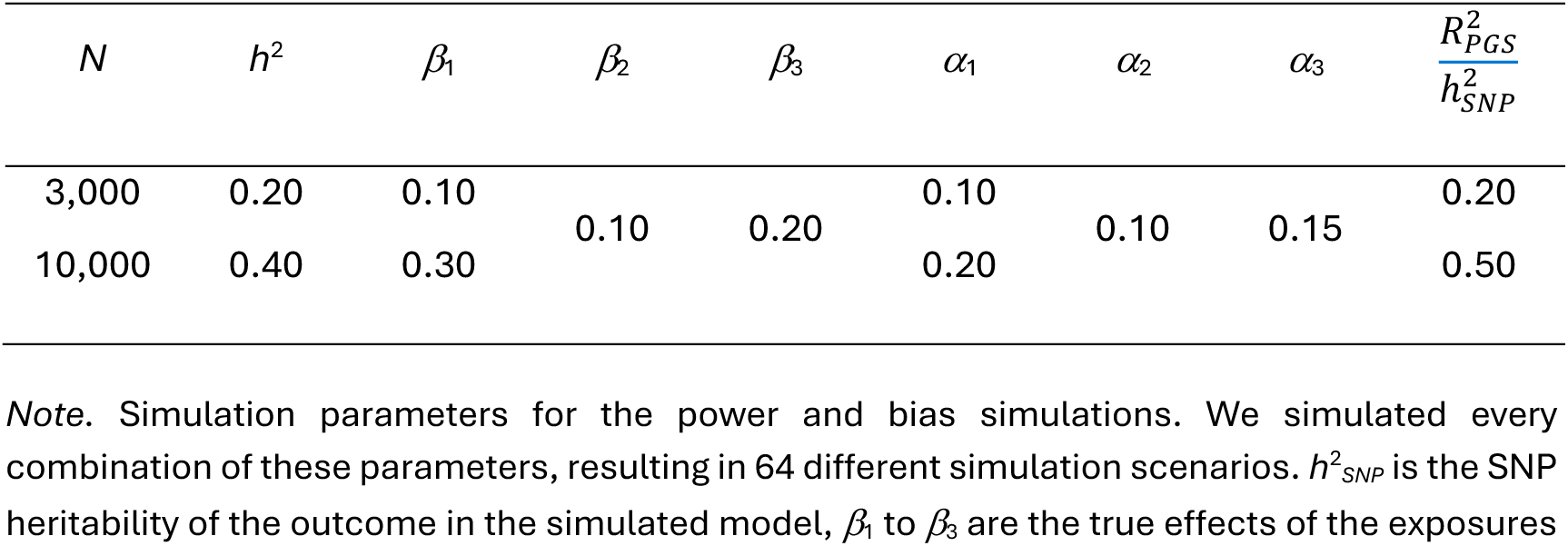
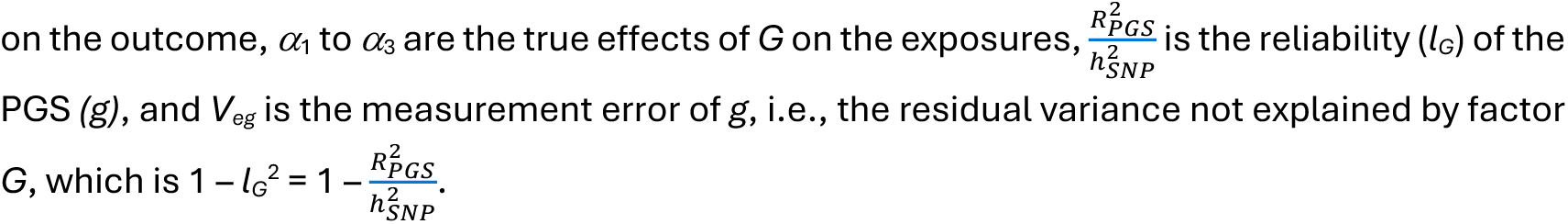
Simulation parameters for power and bias analyses.

To assess type 1 error rates, we used the same parameters for sample size, heritability and measurement error, but restricted to two exposures for simplicity. We set either the *β* or *α* parameters to zero in the (true) theoretical model (i.e., either the effect of an exposure on the outcome is zero, or the effect of *G* on an exposure is zero). For these scenarios, we could therefore assess the type 1 error rate for the exposure-outcome associations (if *β* = 0), or the type 1 error rate for the mediation effects (if either *α*\**β* = 0) or for the genetic confounding estimates (if *α* = 0). *β*_1_ was set to either 0 or 0.30, *β*_2_ was set to 0 or 0.15, *α*_1_ was set to either 0 or 0.10 and *α*_2_ was either 0 or 0.20 for the type 1 error simulations. Each combination of parameters was analysed with 1,000 iterations to assess power, bias or type 1 error, respectively.

We also provide simulations of misspecified models, that is, including causal effects between exposures in the underlying model but omitting the causal effect in the specification of the fitted *Gsens* model (**Table 4**).

**Table 4.**
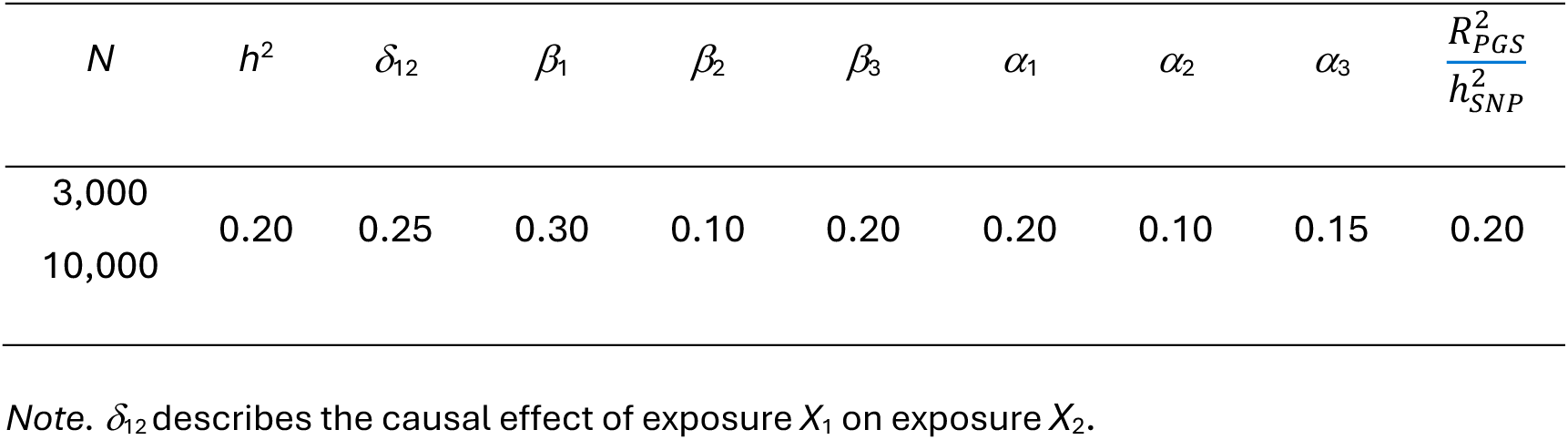
Simulation parameters for misspecified models.

### Sample description

MoBa is a population-based pregnancy cohort study conducted by the Norwegian Institute of Public Health. Participants were recruited from all over Norway from 1999-2008. The women consented to participation in 41% of the pregnancies. The cohort includes approximately 114 500 children, 95 200 mothers and 75 200 fathers. Blood samples were obtained from both parents during pregnancy and from mothers and children (umbilical cord) at birth (53). The current study is based on version 12 of the quality-assured data files released for research in January 2019. The establishment of MoBa and initial data collection was based on a license from the Norwegian Data Protection Agency and approval from The Regional Committees for Medical and Health Research Ethics (REK). The MoBa cohort is now based on regulations related to the Norwegian Health Registry Act. The current study was approved by REK (2016/1702). Furthermore, we used data from the Medical Birth Registry of Norway (MBRN), a national health registry containing information about all births in Norway. In the analytic sample, we use genetic data of 76,207 children (see **Supplementary Table S1** for descriptive statistics of the whole MoBa sample and the analytic sample with genetic data). Phenotypic data for exposures and outcomes was available for 31,476 to 76,207 individuals and data was extracted using the R package *phenotools* v0.4.0 (54) within MoBa’s virtual working environment TSD. As the genotyped MoBa sample contains related individuals (e.g., siblings or cousins), we ran models on the whole genotyped sample and on a subsample of unrelated individuals as sensitivity analyses.

### Measures

#### Exposures

For the first application example, we used birth weight (measured in g and transformed to kg; *N* = 76,207) and the two temperament dimensions ‘Emotionality’ and ‘Activity’ measured by the Emotionality, Activity and Shyness (EAS) Temperament Questionnaire (55,56) (*N* = 42,644–42,674). For the second application example, we used maternal smoking during pregnancy at the start of pregnancy and after the pregnancy was known (*N* = 62,316–64,408; 0 = never, 1 = sometimes, 2 = daily, numerically coded) and maternal pre-pregnancy body mass index (BMI) (*N* = 69,530; kg/m^2^). Maternal and paternal education were mother-rated from 1 (9-year secondary school) to 6 (university, technical college, more than 4 years (master’s degree, medical doctor, PhD)) and were used as numerically coded variables.

#### Outcome

We used an ADHD sum score measured at age 8 years using the Rating Scale for Disruptive Behavior Disorders (RS-DBD; *N* = 31,476; (57), aggregating the subscales of inattention and hyperactivity.

#### Covariates

We used child sex as covariates in all models (48.9% female). For models in example two (intergenerational model), we further adjust for maternal and paternal age at pregnancy.

#### Polygenic score

We calculated a PGS for ADHD based on genome-wide summary statistics (58) using the LDpred2-auto option (59), using an extended set of HapMap3 SNPs and the UK Biobank as LD reference sample. The PGS was standardised to have a mean of zero and a standard deviation of one. Then, we adjusted for population stratification and batch effects by regressing out the first ten principal components of genetic ancestry as well as genotyping batch and array.

### Gsens specifications

We ran *Gsens* v0.1.6 in R v4.1.2 for both applications using complete data with maximum likelihood estimation (default estimator), as missingness is over 50% for the outcome data (∼60% missingness). For the first application example (using birth weight and child temperament as exposures; **Figure 6a**), we assumed a causal effect from birth weight on both temperament dimensions, as suggested in several studies (32) and tested three models for comparison: i) without specifying the causal effect between exposures, ii) specifying a causal effect of birth weight on emotionality, and iii) specifying a causal effect of birth weight on activity. For the second example (using parental education, maternal BMI and smoking during pregnancy as exposures, **Figure 6b**), we did not specify any causal effects between the exposures. Notably, for the second example we also assume no mediation effects of child genotype on ADHD via parental exposures as the exposures precede birth (that is, evocative gene-environment correlations are not plausible). For the heritability estimate, we used the number reported in the original publication (*h*^2^*_SNP_* = 0.14) (58).

**Figure 6.**
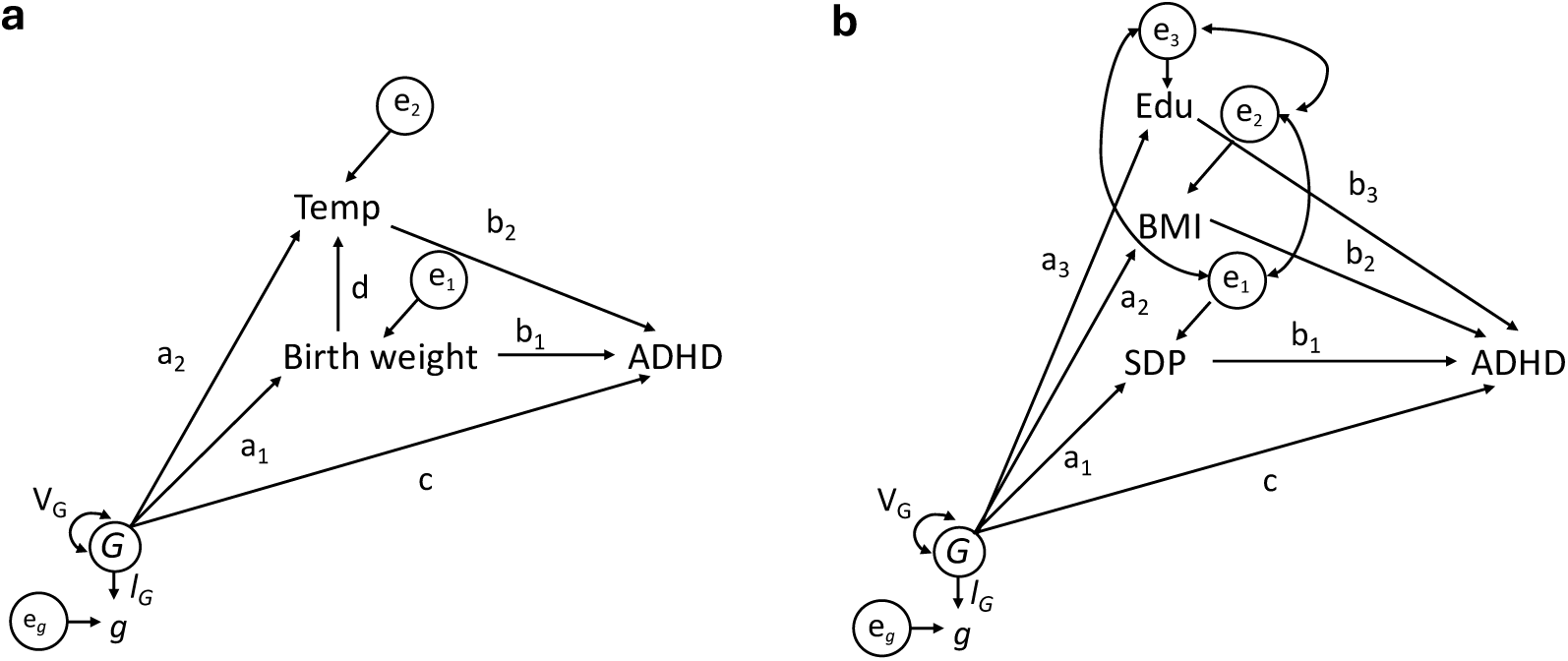
Application examples for Gsens. *Note.* Temp = temperament, BMI = maternal pre-pregnancy BMI, Edu = Parental education, SDP = smoking during pregnancy

## Data and code availability

Data from the Norwegian Mother, Father and Child Cohort Study and the Medical Birth Registry of Norway used in this study are managed by the national health register holders in Norway (Norwegian Institute of public health) and can be made available to researchers, provided approval from the Regional Committees for Medical and Health Research Ethics (REC), compliance with the EU General Data Protection Regulation (GDPR) and approval from the data owners. The consent given by the participants does not open for storage of data on an individual level in repositories or journals. Researchers who want access to datasets for replication should apply through helsedata.no. GWAS summary statistics are publicly available via the Psychiatric Genomics Consortium ([PGC](https://pgc.unc.edu/)). Code for the statistical analyses, the software package and simulation results can be found on [GitHub](https://github.com/LeonardFrach/Gsens), as well as the [pipeline](https://github.com/AndreAllegrini/LDpred2) to compute PGS in MoBa.

## Acknowledgements

The Norwegian Mother, Father and Child Cohort Study is supported by the Norwegian Ministry of Health and Care Services and the Ministry of Education and Research. We are grateful to all the participating families in Norway who take part in this ongoing cohort study. We thank the Norwegian Institute of Public Health (NIPH) for generating high-quality genomic data. This research is part of the HARVEST collaboration, supported by the Research Council of Norway (#229624). We also thank the NORMENT Centre for providing genotype data, funded by the Research Council of Norway (#223273), South East Norway Regional Health Authority and Stiftelsen Kristian Gerhard Jebsen. We further thank the Center for Diabetes Research, the University of Bergen for providing genotype data and performing quality control and imputation of the data funded by the European Research Council (ERC) AdG project SELECTionPREDISPOSED, Stiftelsen Kristian Gerhard Jebsen, Trond Mohn Foundation, the Research Council of Norway, the Novo Nordisk Foundation, the University of Bergen, and the Western Norway Regional Health Authority. This work was performed on the TSD (Tjeneste for Sensitive Data) facilities, owned by the University of Oslo, operated and developed by the TSD service group at the University of Oslo, IT-Department (USIT). The statistical analyses were performed on resources provided by Sigma2—the National Infrastructure for High Performance Computing and Data Storage in Norway. LF and JB-P were supported by the ERC under the European Union’s Horizon 2020 Research and Innovation Programme (grant No. 863981). This work was funded by the European Union’s Horizon Europe Research and Innovation Programme (FAMILY, grant No. 101057529) under the UK government’s Horizon Europe funding guarantee (UK Research and Innovation grant No. 575067).

## Notes

### Competing Interest Statement

The authors have declared no competing interest.

### Author Declarations

The establishment of the Norwegian Mother, Father and Child Cohort (MoBa) study and initial data collection was based on a license from the Norwegian Data Protection Agency and approval from The Regional Committees for Medical and Health Research Ethics (REK). The MoBa cohort is now based on regulations related to the Norwegian Health Registry Act. The current study was approved by REK (2016/1702).

