## Supplements for "Genetic sensitivity analysis: estimating genetic confounding and environmentally mediated genetic effects using multiple exposures"

The aim of this document is to provide derivations related to the quantities referred to in the manuscript. We start with derivations based on two exposures to illustrate the concepts and then generalise to *k* exposures. We present parameters in true (theoretical) models with Greek letters (e.g., $\beta_1$) and parameters in fitted models with Roman letters (e.g., $b_1$). *G* stands for the true genetic factor and *g* is the polygenic score as a measure of *G*. Furthermore, a prime indicates a standardised variable, e.g., the variance of $X_1$ ($V_{X1}$) becomes $V'_{X1}$ when standardised (set to 1).

**Table of contents**

1. *Gsens* equations – general model with multiple exposures
2. Multiple exposures, measurement error in *g* in the true model
3. Multiple exposures – causal effect between exposures
4. Multiple exposures – environmental confounder between exposures
5. Equivalence of equality constraints – heritability vs ratio
6. Amplification bias (collider bias)
7. Supplementary results for the *Gsens* empirical applications in MoBa

### 1. *Gsens* equations – general model with multiple exposures

For illustration of concepts, derivations in this section show a simplified version where *G*, *Y* and the exposures  are measured without error, so the fitted model equals the true model (e.g., if a measured PGS, *g*, captures the true genetic effects). Note that the issue of measurement error in *G*, which we account for in *Gsens*, is addressed in later sections. This section shows that the model allows for a residual covariance between exposures beyond the covariance arising from *G*. A residual covariance between exposures can be present due to a direct relationship (e.g., $X_1 \rightarrow X_2$) or due to common causes not captured by *G* ($X_1 \leftarrow E \rightarrow X_2$).

**Figure S1** The true model

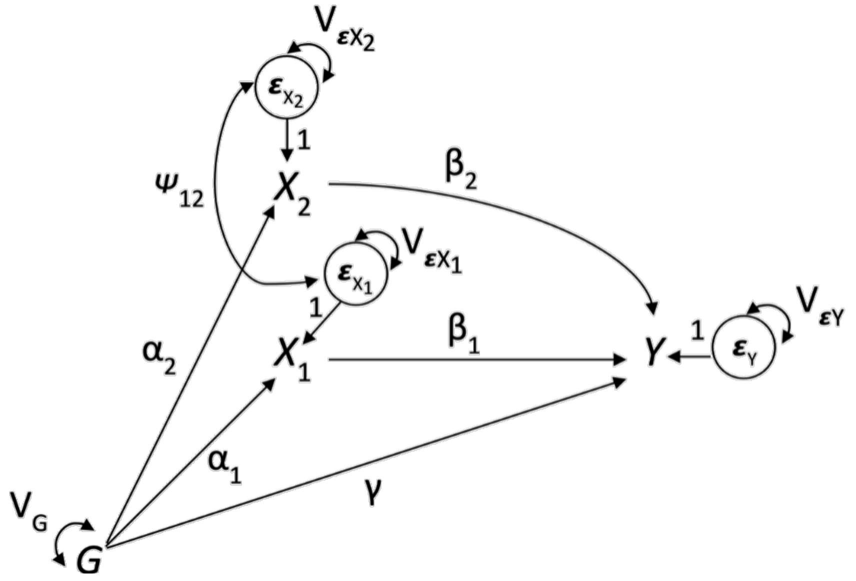


**True model equations**

$$Y = \gamma G + \beta_1 X_1 + \beta_2 X_2 + \varepsilon_Y$$

$$X_1 = \alpha_1 G + \varepsilon_{X_1}$$

$$X_2 = \alpha_2 G + \varepsilon_{X_2}$$

$$G = G$$

**Implied assumptions from the structural model:**

- Error terms for different variables are uncorrelated
    - **Exception**: When there is a common cause *E* with effects on  $X_1$ and $X_2$, or when there is a causal effect of $X_1$ on $X_2$ (or vice versa), then $\text{cov}(\varepsilon_{X1}, \varepsilon_{X2}) \neq 0$
- An error term of a variable is uncorrelated with another variable

- **Exception**: When predictor and outcome are endogenous (e.g. $X_1$ and $Y$), the error term of the predictor ($\varepsilon_{X1}$) is correlated with the outcome variable (Y), which can be seen from path tracing. Thus, Cov($Y$, $\varepsilon_{X1}$) and Cov($Y$, $\varepsilon_{X2}$) ≠ 0

- Furthermore, the covariance terms between exposures and $\varepsilon_Y$ are uncorrelated, as can be seen from path tracing.

**Derive variances in the true model.**

*Variance of Y:*

$$Var(Y) = Cov(Y, Y) = Cov(\gamma G + \beta_1 X_1 + \beta_2 X_2 + \varepsilon_Y, \gamma G + \beta_1 X_1 + \beta_2 X_2 + \varepsilon_Y)$$
$$= Cov(\gamma G, \gamma G) + Cov(\gamma G, \beta_1 X_1) + Cov(\gamma G, \beta_2 X_2) + Cov(\gamma G, \varepsilon_Y) + Cov(\beta_1 X_1, \gamma G) + Cov(\beta_1 X_1, \beta_1 X_1) +$$
$$Cov(\beta_1 X_1, \beta_2 X_2) + Cov(\beta_1 X_1, \varepsilon_Y) + Cov(\beta_2 X_2, \gamma G) + Cov(\beta_2 X_2, \beta_1 X_1) + Cov(\beta_2 X_2, \beta_2 X_2) +$$
$$Cov(\beta_2 X_2, \varepsilon_Y) + Cov(\varepsilon_Y, \gamma G) + Cov(\varepsilon_Y, \beta_1 X_1) + Cov(\varepsilon_Y, \beta_2 X_2) + Cov(\varepsilon_Y, \varepsilon_Y)$$

The residual covariance between exposures $X_1$ and $X_2$ is defined as $\psi_{12}$. As we assume $\varepsilon_Y$ to be uncorrelated to any predictors in the model we get

$$Var(Y) = \gamma^2 V_G + \gamma\beta_1 Cov(G, X_1) + \gamma\beta_2 Cov(G, X_2) + \beta_1\gamma Cov(X_1, G) + \beta_1^2 V_{X_1} + \beta_1\beta_2 Cov(X_1, X_2) +$$
$$\beta_2\gamma Cov(X_2, G) + \beta_2\beta_1 Cov(X_2, X_1) + \beta_2^2 V_{X_2} + V_{\varepsilon_Y}$$
$$= \gamma^2 V_G + 2\gamma\beta_1 Cov(G, X_1) + 2\gamma\beta_2 Cov(G, X_2) + 2\beta_1\beta_2 Cov(X_1, X_2) + \beta_1^2 V_{X_1} + \beta_2^2 V_{X_2} + V_{\varepsilon_Y}$$
$$= \gamma^2 V_G + 2\gamma\beta_1\alpha_1 V_G + 2\gamma\beta_2\alpha_2 V_G + 2\beta_1\beta_2\left[\alpha_1\alpha_2 V_G + \psi_{12}\right] + \beta_1^2 V_{X_1} + \beta_2^2 V_{X_2} + V_{\varepsilon_Y}$$

If standardised, i.e., $V_G$, $V_{X1}$ and $V_{X2}$ = 1, we get the standardised version of $V_Y$ (i.e., $V'_Y$), with $V'_{\varepsilon Y}$ also being standardised and allowing $V'_Y$ to equal 1.

$$V'_Y = \gamma^2 + 2\gamma\alpha_1\beta_1 + 2\gamma\alpha_2\beta_2 + 2\beta_1\beta_2(\alpha_1\alpha_2 + \psi'_{12}) + \beta_1^2 + \beta_2^2 + V'_{\varepsilon_Y}$$

Note that the term $\psi_{12}$ generally reflects a residual covariance between the exposures, e.g., due to a common cause of the exposures (see section 3), whereas $\psi'_{12}$ is the standardised version. If the residual covariance was due to a causal effect from one exposure on the other, this influences the definition of the mediating pathways as explored in misspecification model 1 (section 4).

The variance of $X_1$ is*:*

$$V_{X_1} = Cov(X_1, X_1) = Cov(\alpha_1 G + \varepsilon_{X_1}, \alpha_1 G + \varepsilon_{X_1})$$
$$= \alpha_1^2 V_G + V_{\varepsilon_{X_1}}$$

Similarly, variance of $X_2$ is*:*

$$V_{X_2} = \alpha_2^{\ 2} V_G + V_{\varepsilon_{X_2}}$$

We can replace the variances of $X_1$ and $X_2$ in the final line of the equation for $V_Y$, which becomes

$$Var(Y) = \gamma^2 V_G + 2\gamma\beta_1\alpha_1 V_G + 2\gamma\beta_2\alpha_2 V_G + 2\beta_1\beta_2\left[\alpha_1\alpha_2 V_G + Cov(\varepsilon_{X_1}, \varepsilon_{X_2})\right] + \beta_1^{\ 2}\left(\alpha_1^{\ 2} V_G + V_{\varepsilon_{X_1}}\right) + \beta_2^{\ 2}\left(\alpha_2^{\ 2} V_G + V_{\varepsilon_{X_2}}\right) + V_{\varepsilon_Y}$$

Factoring out $V_G$ from all terms in the variance equation and removing terms that do not include $V_G$, we get the proportion of variance in $Y$ explained by $G$, i.e., the **heritability** ($h^2$)

$$h^2 = \frac{V_G\left(\gamma^2 + 2\gamma\beta_1\alpha_1 + 2\gamma\beta_2\alpha_2 + \beta_1^{\ 2}\alpha_1^{\ 2} + \beta_2^{\ 2}\alpha_2^{\ 2} + 2\beta_1\beta_2\alpha_1\alpha_2\right)}{V_Y}$$

When standardised ($V_G$ and $V_Y = 1$), this heritability expression is

$$h^2 = \gamma^2 + 2\gamma\beta_1\alpha_1 + 2\gamma\beta_2\alpha_2 + \beta_1^{\ 2}\alpha_1^{\ 2} + \beta_2^{\ 2}\alpha_2^{\ 2} + 2\beta_1\beta_2\alpha_1\alpha_2$$

From path tracing, we note that the standardized effect of $G$ on $Y$ is

$$h = \gamma + \alpha_1\beta_1 + \alpha_2\beta_2,$$

thus, this term squared gives us the equal expression:

$$h^2 = \left(\gamma + \alpha_1\beta_1 + \alpha_2\beta_2\right)^2$$

Importantly, heritability of $Y$ was constrained to be a desired external value in the original *Gsens*, and this constraint can still be used in the updated *Gsens* version.

**Derive covariances.**

*Covariance between exposure 1 and the outcome:*

$$\begin{aligned}
Cov(X_1, Y) &= Cov(\alpha_1 G + \varepsilon_{X_1}, \gamma G + \beta_1 X_1 + \beta_2 X_2 + \varepsilon_Y) \\
&= Cov(\alpha_1 G, \gamma G) + Cov(\alpha_1 G, \beta_1 X_1) + Cov(\alpha_1 G, \beta_2 X_2) + Cov(\alpha_1 G, \varepsilon_Y) + \\
&\quad Cov(\varepsilon_{X_1}, \gamma G) + Cov(\varepsilon_{X_1}, \beta_1 X_1) + Cov(\varepsilon_{X_1}, \beta_2 X_2) + Cov(\varepsilon_{X_1}, \varepsilon_Y) \\
&= \alpha_1\gamma V_G + \alpha_1\beta_1 Cov(G, X_1) + \alpha_1\beta_2 Cov(G, X_2) + \beta_1 Cov(\varepsilon_{X_1}, X_1) + \beta_2 Cov(\varepsilon_{X_1}, X_2) \\
&= \alpha_1\gamma V_G + \alpha_1\beta_1 Cov(G, \alpha_1 G + \varepsilon_{X_1}) + \alpha_1\beta_2 Cov(G, \alpha_2 G + \varepsilon_{X_2}) + \\
&\quad \beta_1 Cov(\varepsilon_{X_1}, \alpha_1 G + \varepsilon_{X_1}) + \beta_2 Cov(\varepsilon_{X_1}, \alpha_2 G + \varepsilon_{X_2}) \\
&= \alpha_1\gamma V_G + \alpha_1^{\ 2}\beta_1 V_G + \alpha_1\alpha_2\beta_2 V_G + \beta_1 V_{\varepsilon_{X_1}} + \beta_2\psi_{12}
\end{aligned}$$

Note that if we factor $\beta_1$ from all terms including $\beta_1$, we get

$$\beta_1\left(\alpha_1^2 V_G + V_{\varepsilon_{X_1}}\right)$$

and this term thus decomposes the 'causal effect' $\beta_1$ into two terms, one originating in $G$ and the other originating in the nongenetic part of $X_1$. This way, the covariance can be expressed as

$$Cov(X_1, Y) = \beta_1\left(\alpha_1^2 V_G + V_{\varepsilon_{X_1}}\right) + \alpha_1\gamma V_G + \alpha_1\alpha_2\beta_2 V_G + \beta_2\psi_{12}$$

When standardised (i.e., $V_{X1} = 1$), the covariance between $X_1$ and $Y$ becomes the correlation:

$$r_{X_1Y} = \beta_1 + \alpha_1\gamma + \alpha_1\alpha_2\beta_2 + \beta_2\psi'_{12}$$

Going back to the equation before, we can see that the covariance is equal to the following five terms:

- $\beta_1 V_{\varepsilon X1}$: a term corresponding to the 'causal effect' ($\beta_1$) scaled by the residual variance in $X_1$ ($V_{\varepsilon X1}$), i.e. not explained by predictors of $X_1$. Here this term is what in the direct effect of $X_1$ on $Y$ is not originating in genetic effects and can thus be called the **environmental association** between the exposure and the outcome.
- $\beta_1\alpha_1^2 V_G$: term corresponding to the part of the 'causal effect' of the exposure that is explained by $G$.
- $\alpha_1\gamma V_G + \alpha_1\alpha_2\beta_2 V_{G:}$ two terms corresponding to **genetic confounding** of the association between $X_1$ and $Y$ via $G$:
- $\beta_2\psi_{12}$: backdoor environmental path via the correlated exposure

**Genetic confounding** for any $X_i$-$Y$ association can thus be expressed as

$$C_i = \alpha_i\left(\sum_{j=1, j\neq i}^{k-1} \alpha_j\beta_j + \gamma\right)$$

For the $i^{th}$ exposure and all $k$ exposures.

In our five terms, adding genetic confounding ($\alpha_1\gamma V_G$ + $\alpha_1\alpha_2\beta_2 V_{G)}$ and the part of the 'causal effect' that is due to $G$ ($\beta_1\alpha_1^2 V_{G)}$ provides the **genetic overlap**, i.e. how much of the association between an exposure and an outcome originates in genetic effects. Genetic overlap can be generally expressed as

$$O_i = C_i + \alpha_i^2\beta_i$$

To recapitulate, in our example with two exposures and assuming that the residual covariance between $X_1$ and $X_2$ is null, we can decompose the association between $X_1$ and $Y$ in two ways:

**'Causal effect'** ($\beta_1 V_{\varepsilon X1} + \beta_1\alpha_1^2 V_G$) + **genetic confounding** ($\alpha_1\gamma V_G + \alpha_1\alpha_2\beta_2 V_G$):

or into

**Environmental association** ($\beta_1 V_{\varepsilon X1}$) + **genetic overlap** ($\beta_1\alpha_1^2 V_G + \alpha_1\gamma V_G + \alpha_1\alpha_2\beta_2 V_G$):

The first decomposition ('causal effect' + genetic confounding) was already introduced in the previous *Gsens* publication for one exposure.

**Importantly**, $\beta_1$ (and equivalently $\beta_2$) is the causal effect only in the unrealistic case when no unmeasured nongenetic confounding exists. So beyond our simplified examples without any nongenetic confounders, the term ***residual association*** might be preferred instead.

For multiple exposures, this residual association is similar to the one case exposure but is also adjusted for the covariance between exposures (e.g., $\psi_{ij}$). This means that the total association only equals a strict sum of the residual association + genetic confounding when there is no residual covariance between exposures. It also means that the residual association is not only adjusted for genetic confounding but is also adjusted for the confounding effect due to other exposures in the model. As such, *Gsens* with multiple exposures not only adjust for genetic confounding but also for all the nongenetic component of all exposures explicitly measured in the model. The same applies to the covariance between exposure 2 and the outcome. Practically, measured nongenetic confounders can be additionally accounted for as we show in the application examples. These 'nongenetic' factors (e.g., age and sex) preferably can be regressed out prior to running *Gsens* if they are assumed to be completely unrelated to $G$.

*Covariance between exposure 1 and G:*

$$\begin{aligned} Cov(G, X_1) &= Cov(G, \alpha_1 G + \varepsilon_{X_1}) \\ &= \alpha_1 V_G \end{aligned}$$

When $G$ is standardised, these will be:

$$r_{GX_1} = \alpha_1$$

$$r_{GX_2} = \alpha_2$$

*Covariance between G and Y:*

$$\begin{aligned} Cov(G, Y) &= Cov(G, \gamma G + \beta_1 X_1 + \beta_2 X_2 + \varepsilon_Y) \\ &= \gamma V_G + \beta_1 Cov(G, X_1) + \beta_2 Cov(G, X_2) \\ &= \gamma V_G + \beta_1 \alpha_1 V_G + \beta_2 \alpha_2 V_G \end{aligned}$$

When $G$ is standardised, these will be:

$$r_{GY} = \gamma + \alpha_1 \beta_1 + \alpha_2 \beta_2,$$

which is the square root of the heritability $h^2$.

The association between $G$ and $Y$ comprises the **environmentally mediated genetic effects**, which for two exposures and when $G$ is standardised can be expressed as

$$M_{total} = \alpha_1 \beta_1 + \alpha_2 \beta_2 = r_{GY} - \gamma$$

And for individual mediation paths, this will be

$$M_i = \alpha_i \beta_i$$

---

**Generalisation to *k* exposures**

Expressions below are based on the example with two exposures. Estimates of interest are the covariances (correlations) between the exposures and the outcome.

Using sum notation we have:

**Model equations:**

$$\begin{aligned} Y &= \gamma G + \sum_{i=1}^{k} \beta_i X_i + \varepsilon_Y \\ X_i &= \alpha_i G + \varepsilon_{X_i} \\ G &= G \end{aligned}$$

$$C_i = \alpha_i \left( \sum_{j = 1,\, j \neq i}^{k-1} \alpha_j \beta_j + \gamma \right)$$

For each of $k$ exposures with $i \neq j$, we have:

$$r_{X_iY} = \beta_i V'_{\varepsilon_{Xi}} + \alpha_i\left(\gamma + \sum_{j=1,\, j\neq i}^{k-1} \alpha_j\beta_j\right) + \alpha_i^2\beta_i + \sum_{j=1,\, j\neq i}^{k-1} \beta_j\psi'_{ij}$$

or by joining the genetic and environmental component of the causal effect:

$$r_{X_iY} = \beta_i + \alpha_i\left(\gamma + \sum_{j=1,\, j\neq i}^{k-1} \alpha_j\beta_j\right) + \sum_{j=1,\, j\neq i}^{k-1} \beta_j\psi'_{ij}$$

where $\psi'_{ij}$ describes the residual correlation between exposures $X_i$ and $X_j$. For any exposure $X_i$, there will be $k$–1 residual covariances/correlations with other exposures.

### 2. Multiple exposures, measurement error in *g* in the true model

This is an extension of section 1 to integrate measurement error in *g* (PGS) in the true model to see how estimates would be biased by not accounting for the true measurement error in the fitted model. In a recent review (Pingault et al., 2022), we already presented a set of derivations addressing this issue. Here, we extend the equations to multiple exposures to see how the estimated quantities are **biased** when measurement error in *g* is not adjusted for. We do not model measurement error in *X* and *Y*, but this can be extended further. For simplicity, to highlight the issue of measurement error in *g*, we first present a model without confounders between exposures and the outcome and without any residual correlation between exposures. See sections 3 and 4 for scenarios with correlated exposures.

**Figure S2a** The true model

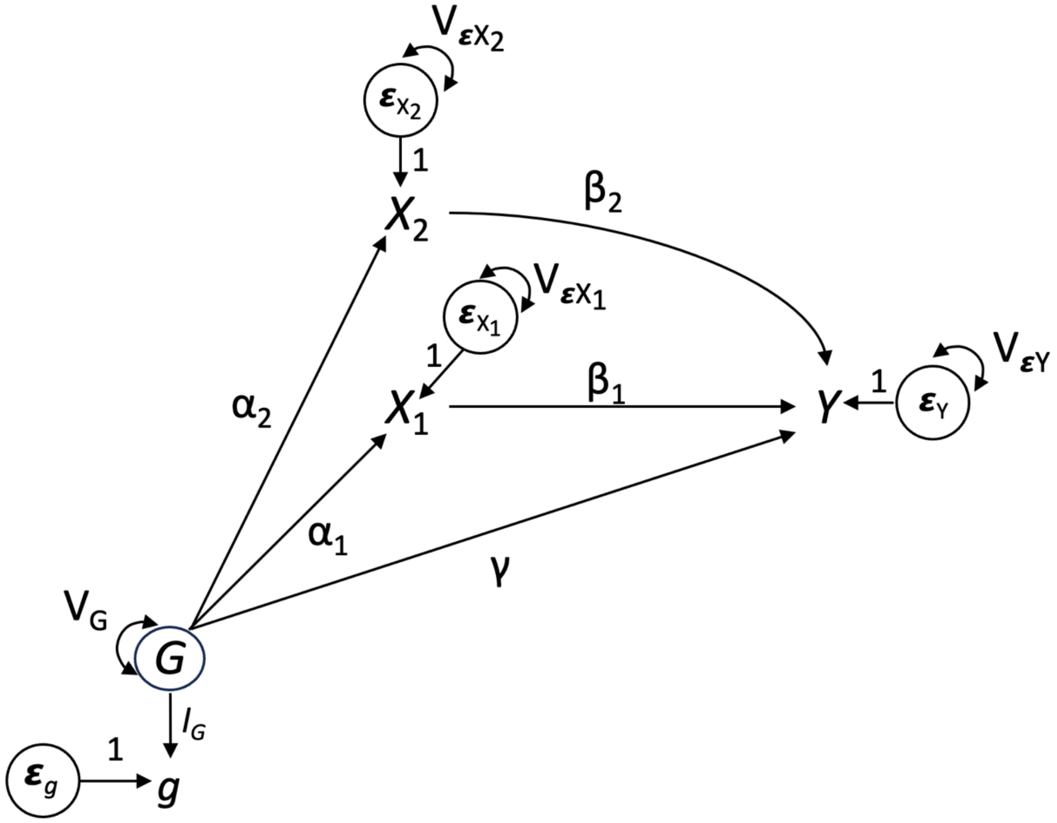


**True model equations**

$$Y = \gamma G + \beta_1 X_1 + \beta_2 X_2 + \varepsilon_Y$$
$$X_1 = \alpha_1 G + \varepsilon_{X_1}$$
$$X_2 = \alpha_2 G + \varepsilon_{X_2}$$
$$g = l_G G + \varepsilon_g$$
$$G = G$$

**Figure S2b** Fitted model using *g* (PGS) – assuming no residual covariance between exposures and outcome, no residual covariance between exposures and perfect measurement of *X*s and *Y*

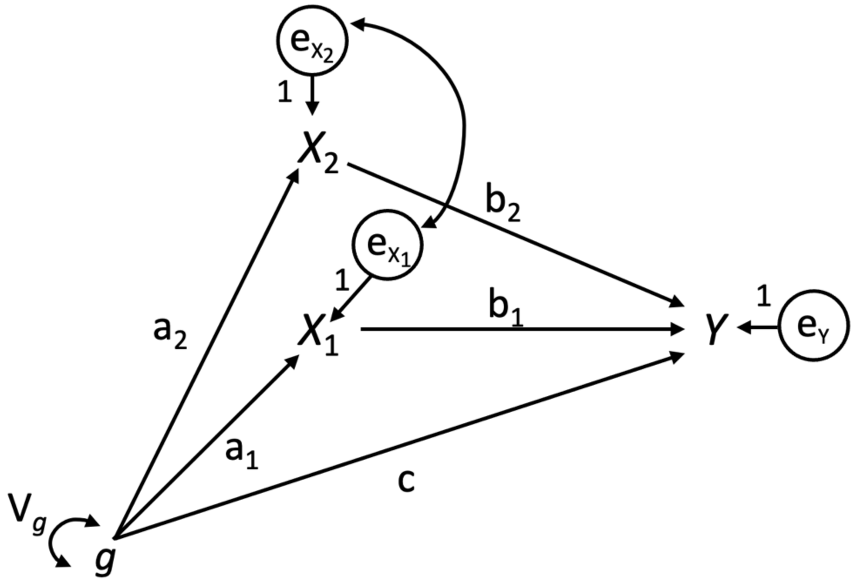


**Fitted model equations**

$$Y = cg + b_1 X_1 + b_2 X_2 + e_Y$$

$$X_1 = a_1 g + e_{X_1}$$

$$X_2 = a_2 g + e_{X_2}$$

$$g = g$$

Note that this fitted model refers to the case where one would just use the PGS. In *Gsens*, we fit a model that explicitly adjusts for the measurement error in *g* (the PGS).

**Step 1. Variances and covariances in the fitted model**

Following similar steps to section 1, we obtain:

$$Var(Y) = c^2 V_g + 2ca_1 b_1 V_g + 2ca_2 b_2 V_g + 2b_1 b_2 a_1 a_2 V_g + b_1^2 V_{X_1} + b_2^2 V_{X_2} + V_{e_Y}$$

$$Var(X_1) = a_1^2 V_g + V_{e_{X_1}}$$

$$Var(X_2) = a_2^2 V_g + V_{e_{X_2}}$$

$$Cov(X_1, Y) = b_1 V_{X_1} + a_1 c V_g + a_1 a_2 b_2 V_g$$

$$Cov(X_2, Y) = b_2 V_{X_2} + a_2 c V_g + a_1 a_2 b_1 V_g$$

$$Cov(g, X_1) = a_1 V_g$$

$$Cov(g, X_2) = a_2 V_g$$

$$Cov(g, Y) = c V_g + a_1 b_1 V_g + a_2 b_2 V_g$$

When $X_1$, $X_2$, $Y$ and $g$ are standardised (all variances equal to 1), the correlations are

$$r_{X_1 Y} = b_1 + a_1 c + a_1 a_2 b_2$$

$$r_{X_2 Y} = b_2 + a_2 c + a_1 a_2 b_1$$

$$r_{gX_1} = a_1$$

$$r_{gX_2} = a_2$$

$$r_{gY} = c + a_1 b_1 + a_2 b_2$$

**Step 2. Express *b* (fitted model) as a function of observed correlations.**

$$b_1 = r_{X_1 Y} - a_1 c - a_1 a_2 b_2 \quad (2.1)$$

$$b_2 = r_{X_2 Y} - a_2 c - a_2 a_1 b_1 \quad (2.2)$$

$$c = r_{gY} - a_1 b_1 - a_2 b_2 \quad (2.3)$$

**Replace *c* in equation (2.1) by equation (2.3) and replace $a_1$ and $a_2$**

$$\begin{aligned} b_1 &= r_{X_1 Y} - r_{gX_1}\left(r_{gY} - r_{gX_1} b_1 - r_{gX_2} b_2\right) - r_{gX_1} r_{gX_2} b_2 \\ &= r_{X_1 Y} - r_{gX_1} r_{gY} + r_{gX_1}^{\ 2} b_1 + r_{gX_1} r_{gX_2} b_2 - r_{gX_1} r_{gX_2} b_2 \\ &= r_{X_1 Y} - r_{gX_1} r_{gY} + r_{gX_1}^{\ 2} b_1 \end{aligned}$$

$$b_1\left(1 - r_{gX_1}^{\ 2}\right) = r_{X_1 Y} - r_{gX_1} r_{gY}$$

So

$$b_1 = \frac{\left(r_{X_1 Y} - r_{gX_1} r_{gY}\right)}{\left(1 - r_{gX_1}^{\ 2}\right)} \quad (2.4)$$

**Replace *c* in equation (2.2) by equation (2.3) and replace *a*₁ and *a*₂**

$$b_2 = r_{X_2Y} - r_{gX_2} r_{gY} + r_{gX_2}{}^2 b_2$$
$$b_2\left(1 - r_{gX_2}{}^2\right) = r_{X_2Y} - r_{gX_2} r_{gY}$$

with the fitted path $b_2$

$$b_2 = \frac{\left(r_{X_2Y} - r_{gX_2} r_{gY}\right)}{\left(1 - r_{gX_2}{}^2\right)} \qquad (2.5)$$

Insert equations for $a_1$, $a_2$, $b_1$ and $b_2$ to define $c$.

$$c = r_{gY} - a_1 b_1 - a_2 b_2$$
$$= r_{gY} - \frac{r_{gX_1} r_{X_1Y} - r_{gX_1}{}^2 r_{gY}}{1 - r_{gX_1}{}^2} - \frac{r_{gX_2} r_{X_2Y} - r_{gX_2}{}^2 r_{gY}}{1 - r_{gX_2}{}^2} \qquad (2.6)$$

**Step 3. Find observed correlations as a function of true betas in the true model.**

Similar to section 1, variances and covariances and their standardised versions are as follows:

$$Var(Y) = \gamma^2 V_G + 2\gamma\beta_1\alpha_1 V_G + 2\gamma\beta_2\alpha_2 V_G + 2\beta_1\beta_2\alpha_1\alpha_2 V_G + \beta_1{}^2 V_{X_1} + \beta_2{}^2 V_{X_2} + V_{\varepsilon_Y}$$

$$Var(X_1) = \alpha_1{}^2 V_G + V_{\varepsilon_{X_1}}$$
$$Var(X_2) = \alpha_2{}^2 V_G + V_{\varepsilon_{X_2}}$$

When standardised:

$$Var'(X_1) = \alpha_1{}^2 + V'_{\varepsilon_{X_1}} = 1$$
$$Var'(X_2) = \alpha_2{}^2 + V'_{\varepsilon_{X_2}} = 1$$

**Derive covariances in the true model.**

$$Cov(X_1, g) = l_G \alpha_1 V_G$$
$$Cov(X_2, g) = l_G \alpha_2 V_G$$

Which when standardised are:

$$r_{GX_1} = l_G\alpha_1$$

$$r_{GX_2} = l_G\alpha_2$$

*Covariance between g and the outcome:*

$$Cov(g, Y) = l_G\gamma V_G + l_G\alpha_1\beta_1 V_G + l_G\alpha_2\beta_2 V_G$$

$$r_{gY} = l_G(\gamma + \alpha_1\beta_1 + \alpha_2\beta_2)$$

→ correlation/covariance between *g* (PGS) and the outcome is scaled by $l_G$

*Covariance and correlations between exposures and the outcome:*

$$Cov(X_1, Y) = \beta_1 V_{X_1} + \alpha_1\gamma V_G + \alpha_1\alpha_2\beta_2 V_G$$

$$r_{X_1Y} = \beta_1 + \alpha_1\gamma + \alpha_1\alpha_2\beta_2$$

$$Cov(X_2, Y) = \beta_2 V_{X_2} + \alpha_2\gamma V_G + \alpha_2\alpha_1\beta_1 V_G$$

$$r_{X_2Y} = \beta_2 + \alpha_2\gamma + \alpha_1\alpha_2\beta_1$$

**Step 4. Find fitted paths as a function of the true paths.**

Again, similar to section 1, we get the following:

$$a_1 = r_{gX_1} = l_G\alpha_1$$

$$a_2 = r_{gX_2} = l_G\alpha_2$$

$$b_1 = \frac{r_{X_1Y} - r_{gX_1}r_{gY}}{1 - r_{gX_1}^{\;2}}$$

$$= \beta_1 + \frac{\left(\alpha_1\gamma + \alpha_1\alpha_2\beta_2\right)\left(1 - l_G^{\,2}\right)}{1 - l_G^{\,2}\alpha_1^{\,2}}$$

Therefore, the fitted $b_1$ path consists of the true $\beta_1$ path + a bias term

$$Bias_{b_1} = \frac{\left(\alpha_1\gamma + \alpha_1\alpha_2\beta_2\right)\left(1 - l_G^{\,2}\right)}{1 - l_G^{\,2}\alpha_1^{\,2}}$$

This includes residual genetic confounding, scaled by the measurement error ($1 - l_G^2$). If there is no measurement error (i.e., $l_G^2 = 1$), the numerator becomes null and thus the bias term becomes 0.

Similarly, for $b_2$, we get

$$b_2 = \beta_2 + \frac{\left(\alpha_2\gamma + \alpha_1\alpha_2\beta_1\right)\left(1 - l_G{}^2\right)}{1 - l_G{}^2\alpha_2{}^2}$$

and

$$Bias_{b_2} = \frac{\left(\alpha_2\gamma + \alpha_1\alpha_2\beta_1\right)\left(1 - l_G{}^2\right)}{1 - l_G{}^2\alpha_2{}^2}$$

Lastly, we can insert these terms in the expression of path *c* that we had:

$$\begin{aligned} c &= r_{gY} - a_1 b_1 - a_2 b_2 \\ &= l_G\left(\gamma + \alpha_1\beta_1 + \alpha_2\beta_2\right) - l_G\alpha_1\beta_1 - l_G\alpha_1 Bias_{b_1} - l_G\alpha_2\beta_2 - l_G\alpha_2 Bias_{b_2} \\ &= l_G\gamma - l_G\alpha_1 Bias_{b_1} - l_G\alpha_2 Bias_{b_2} \end{aligned}$$

We can see that, whereas $a_1$ and $a_2$ are only biased by being scaled by $l_G$, paths $b_1$ and $b_2$ include a bias term reflecting residual genetic confounding. Consequently, path *c,* in addition to including the true direct effect $\gamma$ scaled by $l_G$, includes (negative) bias from both paths $b_1$ and $b_2$. For path *c* again, if there is no measurement error ($l_G = 1$), both bias terms from $b_1$ and $b_2$ become zero and $c = \gamma$.

Bias in $b_1$ and $b_2$ would affect all other quantities too, that is, environmentally mediated genetic effects, genetic confounding and genetic overlap.

For the **total mediation**

$$M_G = \sum_{j=1}^{k} a_j b_j = a_1 b_1 + a_2 b_2$$

we get

$$M_G = l_G\left(\alpha_1\beta_1 + \alpha_2\beta_2\right) + l_G\alpha_1 Bias_{b_1} + l_G\alpha_2 Bias_{b_2}$$

We can see that the total mediation is biased, as it contains the true mediation scaled by $l_G$ (downward bias) but also the two additional bias terms of $b_1$ and $b_2$.

Similarly, for the **genetic confounding** estimates

$$C_i = a_i\left(\sum_{j=1,\ j\neq i}^{k} a_j b_j\right) + c$$

We get

$$\begin{aligned} C_1 &= a_1(a_2 b_2 + c) \\ &= l_G^2 \alpha_1 \alpha_2 \beta_2 + l_G^2 \alpha_1 \alpha_2 Bias_{b_2} + l_G \alpha_1 \left( l_G \gamma - l_G \alpha_1 Bias_{b_1} - l_G \alpha_2 Bias_{b_2} \right) \\ &= l_G^2 \left( \alpha_1 \alpha_2 \beta_2 + \alpha_1 \gamma \right) - l_G^2 \alpha_1^2 Bias_{b_1} \end{aligned}$$

and

$$C_2 = l_G^2 \left( \alpha_2 \alpha_1 \beta_1 + \alpha_2 \gamma \right) - l_G^2 \alpha_2^2 Bias_{b_2}$$

These genetic confounding estimates contain the true genetic confounding terms scaled by the reliability of the polygenic score ($l_G^2$) and an additional bias term. In general, as $l_G$ will be smaller than 1, the genetic confounding paths will often be underestimated but also depending on the bias of $b_i$. Again, if $l_G = 1$, there is no bias and the estimate of genetic confounding equals the true genetic confounding.

Finally, for the **genetic overlap**

$$O_i = C_i + a_i^2 b_i$$

We get

$$\begin{aligned} O_1 &= C_1 + l_G^2 \alpha_1^2 \beta_1 + l_G^2 \alpha_1^2 Bias_{b_1} \\ &= l_G^2 \left( \alpha_1 \alpha_2 \beta_2 + \alpha_1 \gamma \right) - l_G^2 \alpha_1^2 Bias_{b_1} + l_G^2 \alpha_1^2 \beta_1 + l_G^2 \alpha_1^2 Bias_{b_1} \\ &= l_G^2 \left( \alpha_1 \alpha_2 \beta_2 + \alpha_1 \gamma + \alpha_1^2 \beta_1 \right) + l_G^2 \alpha_1^2 \beta_1 \end{aligned}$$

and

$$O_2 = l_G^2 \left( \alpha_2 \alpha_1 \beta_1 + \alpha_2 \gamma + \alpha_2^2 \beta_2 \right) + l_G^2 \alpha_2^2 \beta_2$$

Here, the genetic overlap $O_i$ will be scaled by $l_G^2$ but should not be influenced by the bias in $b_i$.

### 3. Causal effect of $X_1$ on $X_2$ in the true model

In this section, we show how the fitted *Gsens* model is affected if there is an unmodelled causal relation between exposures, which exists in the true theoretical model. For simplicity, this section assumes no measurement error in *Y* and in the exposures.

**Figure S3a** The true model

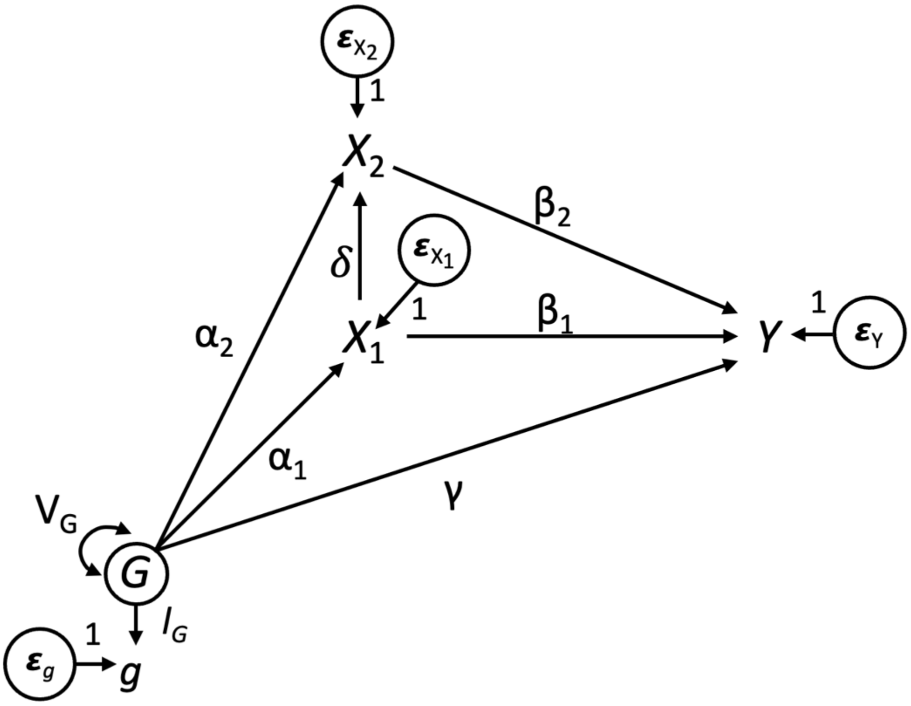


**True model equations:**

$$Y = \gamma G + \beta_1 X_1 + \beta_2 X_2 + \varepsilon_Y$$

$$X_1 = \alpha_1 G + \varepsilon_{X_1}$$

$$X_2 = \alpha_2 G + \delta X_1 + \varepsilon_{X_2}$$

$$g = l_G G + \varepsilon_g$$

$$G = G$$

**Additional assumptions:**

- When there is a causal effect of $X_1$ on $X_2$ (or vice versa), then cov($\varepsilon_{X1}$, $X_2$) or cov($\varepsilon_{X2}$, $X_1$) ≠ 0, respectively, if these causal relations are not taken into account.

**Figure S3b** The fitted model

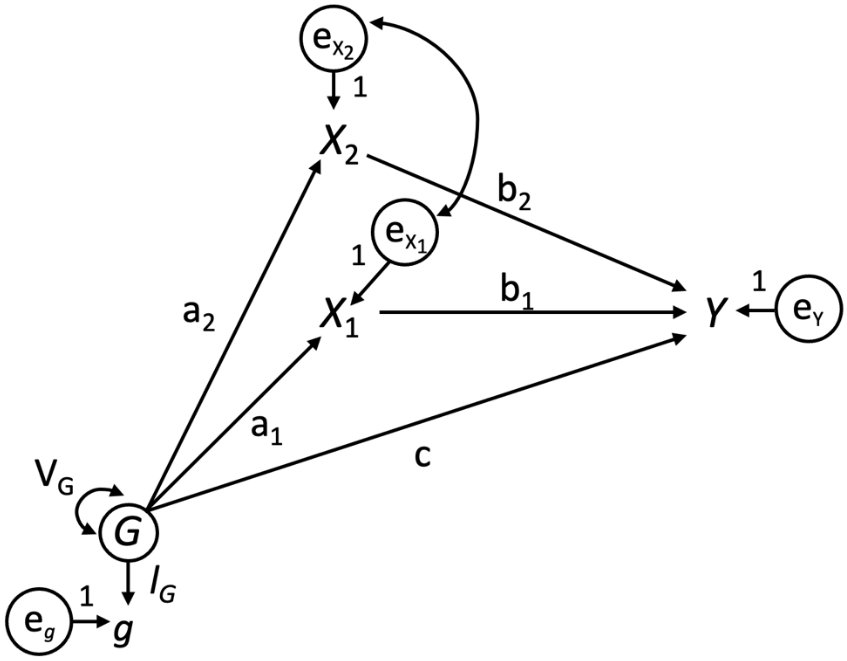


**Fitted model equations** (allowing for residual correlation between exposures):

$$Y = cG + b_1X_1 + b_2X_2 + e_Y$$

$$X_1 = a_1G + e_{X_1}$$

$$X_2 = a_2G + e_{X_2}$$

$$g = l_GG + e_g$$

$$G = G$$

In the fitted model, express the parameters as a function of the observed correlations:

Paths $a_1$ and $a_2$

$$Cov(X_1, G) = Cov(a_1G + e_{X_1}, G) = a_1V_G$$

$$Cov(X_2, G) = Cov(a_2G + e_{X_2}, G) = a_2V_G$$

Standardised:

$$r_{X_1G} = a_1$$

$$r_{X_2G} = a_2$$

Paths $b_1$ and $b_2$

$$Cov(X_1, Y) = b_1 V_{X_1} + a_1 c V_G + a_1 b_2 a_2 V_G + b_2 Cov(e_{X_1}, e_{X_2})$$

Standardised

$$r_{X_1 Y} = b_1 + a_1 c + a_1 a_2 b_2 + b_2 s'_{12}$$

As mentioned in section 1, the last term $s'_{12}$ should correspond to any source of (standardised) covariance (i.e., correlation) between $X_1$ and $X_2$ not due to $G$, for example our misspecified model with causal effect $\delta_{X1}$ of $X_1$ on $X_2$. So, although the causal relationship between $X_1$ and $X_2$ is not explicitly specified in the structural fitted model, it is still modelled via the residual covariance term.

In the fitted model, $X_1$ and $X_2$ both include the additional covariance term in the same way so:

$$r_{X_2 Y} = b_2 + a_2 c + a_1 a_2 b_1 + b_1 s'_{12}$$

**Let us derive *s'*12**

We have

$$Cov(X_1, X_2) = a_1 a_2 V_G + Cov(e_{X_1}, e_{X_2})$$

Standardised:

$$r_{X_1 X_2} = a_1 a_2 + s'_{12}$$

$$s'_{12} = r_{X_1 X_2} - r_{X_1 G} r_{X_2 G}$$

Intuitively, this is the observed correlation minus the part generated by $G$.

Path $c$

$$Cov(G, Y) = cV_G + b_1 a_1 V_G + b_2 a_2 V_G$$

Standardised:

$$r_{GY} = c + a_1 b_1 + a_2 b_2$$

Interestingly, the residual correlation between $e_{X1}$ and $e_{X2}$ does not play a here, reinforcing the notion that $r_{GY}$ is decomposed into the direct effect of $G$ and the indirect effects of $G$ via $X_1$ and $X_2$ without additional paths via the residual correlation.

Express $a_1$, $a_2$, $b_1$, $b_2$, $c$ as a function of correlations:

$$a_1 = r_{X_1G}$$

$$a_2 = r_{X_2G}$$

$$s'_{12} = r_{X_1X_2} - r_{X_1G}r_{X_2G}$$

$$b_1 = r_{X_1Y} - a_1c - a_1a_2b_2 - b_2s'_{12} \quad (3.1)$$

$$b_2 = r_{X_2Y} - a_2c - a_2a_1b_1 - b_1s'_{12} \quad (3.2)$$

$$c = r_{GY} - a_1b_1 - a_2b_2 \quad (3.3)$$

**Replace *c* in equation (3.1) by equation (3.3)**

$$b_1 = r_{X_1Y} - a_1c - a_1a_2b_2 - b_2s'_{12}$$

$$= r_{X_1Y} - a_1r_{GY} + a_1^2b_1 - b_2s'_{12}$$

$$b_1(1 - a_1^2) = r_{X_1Y} - a_1r_{GY} - b_2s'_{12} \quad (3.4)$$

Which gives us

$$b_1 = \frac{(r_{X_1Y} - a_1r_{GY} - b_2s'_{12})}{(1 - a_1^2)}$$

The correlation of the errors complexifies the equation as, without it, $b_2$ is eliminated right away leading to a simple expression of $b_1$.

**Equivalently, replace *c* in equation (3.2) by equation (3.3)**

$$b_2 = \frac{(r_{X_2Y} - a_2r_{GY} - b_1s'_{12})}{(1 - a_2^2)} \quad (3.5)$$

**Replace $b_2$ in equation (3.4) by equation (3.5)**

$$b_1(1-a_1^2) = r_{X_1Y} - a_1 r_{GY} - \frac{s'_{12}(r_{X_2Y} - a_2 r_{GY} - b_1 s'_{12})}{(1-a_2^2)}$$

$$b_1(1-a_1^2)(1-a_2^2) = r_{X_1Y}(1-a_2^2) - a_1 r_{GY}(1-a_2^2) - s'_{12}(r_{X_2Y} - a_2 r_{GY} - b_1 s'_{12})$$

$$b_1\left[(1-a_1^2)(1-a_2^2) - s'^2_{12}\right] = r_{X_1Y} - a_2^2 r_{X_1Y} - a_1 r_{GY} + a_2^2 a_1 r_{GY} - s'_{12} r_{X_2Y} + s'_{12} a_2 r_{GY}$$

Replace

$$a_1 = r_{X_1G}$$

$$a_2 = r_{X_2G}$$

$$s'_{12} = r_{X_1X_2} - r_{X_1G} r_{X_2G}$$

Left part:

$$b_1\left[(1-r_{X_1G}^2)(1-r_{X_2G}^2) - (r_{X_1X_2} - r_{X_1G} r_{X_2G})^2\right]$$
$$= b_1\left(1 - r_{X_2G}^2 - r_{X_1G}^2 - r_{X_1X_2}^2 + 2 r_{X_1X_2} r_{X_1G} r_{X_2G}\right)$$

Right part:

$$r_{X_1Y} - r_{X_2G}^2 r_{X_1Y} - r_{X_1G} r_{GY} + r_{X_2G}^2 r_{X_1G} r_{GY} - (r_{X_1X_2} - r_{X_1G} r_{X_2G}) r_{X_2Y} + (r_{X_1X_2} - r_{X_1G} r_{X_2G}) r_{X_2G} r_{GY}$$
$$= r_{X_1Y} - r_{X_2G}^2 r_{X_1Y} - r_{X_1G} r_{GY} - r_{X_1X_2} r_{X_2Y} + r_{X_1G} r_{X_2G} r_{X_2Y} + r_{X_1X_2} r_{X_2G} r_{GY}$$

**In the true model, express the correlation as a function of betas.**

Variances needed:

$$Var(X_1) = \alpha_1^2 V_G + V_{\varepsilon_{X_1}}$$

$$Var(X_2) = Cov(\alpha_2 G + \delta X_1 + \varepsilon_{X_2}, \alpha_2 G + \delta X_1 + \varepsilon_{X_2})$$
$$= \alpha_2^2 V_G + 2\alpha_2 \alpha_1 \delta V_G + \delta^2 V_{X_1} + V_{\varepsilon_{X_2}}$$

which is consistent with path tracing rules and is no longer symmetrical with $X_1$.

**Covariances and correlations:**

$$Cov(X_1, G) = Cov(\alpha_1 G + \varepsilon_{X_1}, G) = \alpha_1 V_G$$

$$r_{X_1G} = \alpha_1$$

and

$$Cov(X_2, G) = \alpha_2 V_G + \delta\alpha_1 V_G$$

$$r_{X_2 G} = \alpha_2 + \alpha_1\delta ,$$

which differs from the fitted model.

*Covariance and correlation between exposures*

$$\begin{aligned} Cov\left(X_1, X_2\right) &= Cov\left(\alpha_1 G + \varepsilon_{X_1}, \alpha_2 G + \delta X_1 + \varepsilon_{X_2}\right) \\ &= \alpha_1\alpha_2 V_G + \alpha_1\delta Cov\left(G, X_1\right) + \delta\, Cov\left(\varepsilon_{X_1}, X_1\right) \\ &= \alpha_1\alpha_2 V_G + \delta\left(\alpha_1^{\,2} V_G + V_{\varepsilon_{X_1}}\right) \\ &= \alpha_1\alpha_2 V_G + \delta V_{X_1} \end{aligned}$$

Standardised:

$$r_{X_1 X_2} = \delta + \alpha_1\alpha_2$$

*Covariance and correlation between exposures and Y*

$$\begin{aligned} Cov(X_1, Y) &= Cov(\alpha_1 G + \varepsilon_{X_1}, \gamma G + \beta_1 X_1 + \beta_2 X_2 + \varepsilon_Y) \\ &= Cov(\alpha_1 G, \gamma G) + Cov(\alpha_1 G, \beta_1 X_1) + Cov(\alpha_1 G, \beta_2 X_2) + \\ &\quad Cov(\alpha_1 G, \varepsilon_Y) + Cov(\varepsilon_{X_1}, \gamma G) + Cov(\varepsilon_{X_1}, \beta_1 X_1) + \\ &\quad Cov(\varepsilon_{X_1}, \beta_2 X_2) + Cov(\varepsilon_{X_1}, \varepsilon_Y) \\ &= \alpha_1\gamma V_G + \alpha_1\beta_1\alpha_1 V_G + \alpha_1\beta_2(\alpha_2 V_G + \delta\alpha_1 V_G) + \\ &\quad \beta_1 Cov(\varepsilon_{X_1}, \alpha_1 G + \varepsilon_{X_1}) + \beta_2 Cov(\varepsilon_{X_1}, \alpha_2 G + \delta X_1 + \varepsilon_{X_2}) \\ &= \alpha_1\gamma V_G + \alpha_1^{\,2}\beta_1 V_G + \alpha_1\beta_2\alpha_2 V_G + \alpha_1^{\,2}\beta_2\delta V_G + \beta_1 V_{\varepsilon_{X_1}} + \delta\beta_2 V_{\varepsilon_{X_1}} \end{aligned}$$

Standardised and reordered:

$$\begin{aligned} r_{X_1 Y} &= \beta_1\left(\alpha_1^{\,2} + V_{\varepsilon_{X_1}}\right) + \alpha_1\gamma + \alpha_1\beta_2\alpha_2 + \delta\beta_2\left(\alpha_1^{\,2} + V_{\varepsilon_{X_1}}\right) \\ &= \beta_1 + \alpha_1\gamma + \alpha_1\alpha_2\beta_2 + \delta\beta_2 \end{aligned}$$

For the $X_2$-$Y$ association, we get:

$$Cov(X_2, Y) = Cov(\alpha_2 G + \delta X_1 + \varepsilon_{X_2}, \gamma G + \beta_1 X_1 + \beta_2 X_2 + \varepsilon_Y)$$

$$= Cov(\alpha_2 G, \gamma G) + Cov(\alpha_2 G, \beta_1 X_1) + Cov(\alpha_2 G, \beta_2 X_2) +$$

$$Cov(\delta X_1, \gamma G) + Cov(\delta X_1, \beta_1 X_1) + Cov(\delta X_1, \beta_2 X_2) + Cov(\varepsilon_{X_2}, \beta_2 X_2)$$

$$= \alpha_2 \gamma V_G + \alpha_2 \alpha_1 \beta_1 V_G + \alpha_2^2 \beta_2 V_G + 2\alpha_2 \alpha_1 \delta \beta_2 V_G + \delta \alpha_1 \gamma V_G + \delta \beta_1 V_{X_1} +$$

$$\delta^2 \beta_2 V_{X_1} + \beta_2 V_{\varepsilon_{X_2}}$$

Some components add up to the causal effect $\beta_2$. Factoring $\beta_2$, we get:

$$Cov(X_2, Y) = \beta_2(\alpha_2^2 V_G + 2\alpha_2 \alpha_1 \delta V_G + \delta^2 V_{X_1} + V_{\varepsilon_{X_2}}) + \alpha_2 \gamma V_G + \alpha_2 \alpha_1 \beta_1 V_G + \delta \alpha_1 \gamma V_G + \delta \beta_1 V_{X_1}$$

Finally, we have

$$Cov(X_2, Y) = \beta_2 V_{X_2} + \alpha_2 \gamma V_G + \alpha_2 \alpha_1 \beta_1 V_G + \delta \alpha_1 \gamma V_G + \delta \beta_1 V_{X_1}$$

and the standardised version

$$r_{X_2 Y} = \beta_2 + \alpha_2 \gamma + \alpha_2 \alpha_1 \beta_1 + \delta \alpha_1 \gamma + \delta \beta_1 ,$$

which is consistent with path tracing.

*Covariance between G and Y*

$$Cov(G, Y) = \gamma V_G + \beta_1 \alpha_1 V_G + \beta_2 \alpha_2 V_G + \beta_2 \delta \alpha_1 V_G$$

Standardised version

$$r_{GY} = \gamma + \alpha_1 \beta_1 + \alpha_2 \beta_2 + \alpha_1 \delta \beta_2$$

So, the correlation is equal to the direct effect + the mediated effects via $X_1$ & $X_2$ including via $\delta$.

**Express fitted betas as function of true betas.**

From above, we have the following results for the fitted model:

$$a_1 = r_{X_1 G}$$

$$a_2 = r_{X_2 G}$$

$$s'_{12} = r_{X_1 X_2} - r_{X_1 G} r_{X_2 G}$$

And the expression for $b_1$:

$$b_1\left(1 - r_{X_2G}^2 - r_{X_1G}^2 - r_{X_1X_2}^2 + 2\, r_{X_1X_2} r_{X_1G} r_{X_2G}\right) = r_{X_1Y} - r_{GX_2}^2 r_{X_1Y} - r_{GX_1} r_{GY} - r_{X_1X_2} r_{X_2Y} + r_{GX_1} r_{GX_2} r_{X_2Y} + r_{GX_2} r_{GY} r_{X_1X_2}$$

As well as, for the true model

$$r_{X_1G} = \alpha_1$$

$$r_{X_2G} = \alpha_2 + \alpha_1\delta$$

$$r_{X_1X_2} = \delta + \alpha_1\alpha_2$$

$$r_{X_1Y} = \beta_1 + \alpha_1\gamma + \alpha_1\alpha_2\beta_2 + \delta\beta_2$$

$$r_{X_2Y} = \beta_2 + \alpha_2\gamma + \alpha_2\alpha_1\beta_1 + \delta\alpha_1\gamma + \delta\beta_1$$

$$r_{GY} = \gamma + \alpha_1\beta_1 + \alpha_2\beta_2 + \alpha_1\delta\beta_2$$

We have:

$$a_1 = r_{X_1G} = \alpha_1,$$

showing that $a_1$ is an **unbiased** estimate of $\alpha_1$.
In contrast, we have

$$a_2 = r_{X_2G} = \alpha_2 + \alpha_1\delta\ ,$$

showing that $a_2$ is a **biased** estimate of the true path $\alpha_2$ (over-/ or underestimated depending on the sign of $\alpha_1$ and $\delta$).

For $b_1$, we have for the left side of the equation:

$$b_1\left(1 - r_{X_2G}^2 - r_{X_1G}^2 - r_{X_1X_2}^2 + 2\, r_{X_1X_2} r_{X_1G} r_{X_2G}\right)$$
$$= b_1\left[1 - \alpha_2^2 - 2\alpha_2\alpha_1\delta - \alpha_1^2\delta^2 - \alpha_1^2 - \delta^2 - 2\delta\alpha_1\alpha_2 - \alpha_1^2\alpha_2^2 + 2\,\alpha_1\delta\alpha_2 + 2\,\alpha_1^2\delta^2 + 2\,\alpha_1^2\alpha_2^2 + 2\,\alpha_1^3\alpha_2\delta)\right]$$

Reordered, this is

$$b_1[1 - \alpha_1^2 - \alpha_2^2 - \delta^2 + \alpha_1^2\alpha_2^2 + \alpha_1^2\delta^2 - 2\,\delta\alpha_1\alpha_2 + 2\,\alpha_1^3\alpha_2\delta]$$

For the right side of the equation, we had

$$= r_{X_1Y} - r_{GX_2}^2 r_{X_1Y} - r_{GX_1} r_{GY} - r_{X_1X_2} r_{X_2Y} + r_{GX_1} r_{GX_2} r_{X_2Y} + r_{GX_2} r_{GY} r_{X_1X_2}$$

Inserting the true values for the correlations, we get the following 6 lines corresponding to the 6 terms:

$$(i)\,\beta_1 + \alpha_1\gamma + \alpha_1\alpha_2\beta_2 + \delta\beta_2$$
$$(ii) - \left[(\alpha_2 + \alpha_1\delta)^2 (\beta_1 + \alpha_1\gamma + \alpha_1\alpha_2\beta_2 + \delta\beta_2)\right]$$
$$(iii) - \alpha_1(\gamma + \alpha_1\beta_1 + \alpha_2\beta_2 + \alpha_1\delta\beta_2)$$
$$(iv) - \left[(\delta + \alpha_1\alpha_2)(\beta_2 + \alpha_2\gamma + \alpha_2\alpha_1\beta_1 + \delta\alpha_1\gamma + \delta\beta_1)\right]$$
$$(v) + \alpha_1(\alpha_2 + \alpha_1\delta)(\beta_2 + \alpha_2\gamma + \alpha_2\alpha_1\beta_1 + \delta\alpha_1\gamma + \delta\beta_1)$$
$$(vi) + (\alpha_2 + \alpha_1\delta)(\gamma + \alpha_1\beta_1 + \alpha_2\beta_2 + \alpha_1\delta\beta_2)(\delta + \alpha_1\alpha_2)$$

$$= (i)\,\beta_1 + \alpha_1\gamma + \alpha_1\alpha_2\beta_2 + \delta\beta_2$$
$$(ii) - \left[(\alpha_2^2 + 2\alpha_2\alpha_1\delta + \alpha_1^2\delta^2)(\beta_1 + \alpha_1\gamma + \alpha_1\alpha_2\beta_2 + \delta\beta_2)\right]$$
$$(iii) - \alpha_1\gamma - \alpha_1^2\beta_1 - \alpha_1\alpha_2\beta_2 - \alpha_1^2\delta\beta_2$$
$$(iv) - \left[\delta\beta_2 + \alpha_2\delta\gamma + \alpha_2\alpha_1\delta\beta_1 + \delta^2\alpha_1\gamma + \delta^2\beta_1 + \alpha_1\alpha_2\beta_2 + \alpha_1\alpha_2^2\gamma + \alpha_1^2\alpha_2^2\beta_1 + \alpha_1^2\alpha_2\delta\gamma + \alpha_1\alpha_2\delta\beta_1\right]$$
$$(v) + \left[(\alpha_1\alpha_2 + \alpha_1^2\delta)(\beta_2 + \alpha_2\gamma + \alpha_2\alpha_1\beta_1 + \delta\alpha_1\gamma + \delta\beta_1)\right]$$
$$(vi) + \left[(\alpha_2\delta + \alpha_1\alpha_2^2 + \alpha_1\delta^2 + \alpha_1^2\alpha_2\delta)(\gamma + \alpha_1\beta_1 + \alpha_2\beta_2 + \alpha_1\delta\beta_2)\right]$$

Solving this and combining with the left part of the equation of the estimated path $b_1$, we get

$$b_1\left(1 - \alpha_1^2 - \alpha_2^2 - \delta^2 + \alpha_1^2\alpha_2^2 + \alpha_1^2\delta^2 - 2\alpha_1\alpha_2\delta + 2\alpha_1^3\alpha_2\delta\right)$$
$$= \beta_1\left(1 - \alpha_1^2 - \alpha_2^2 - \delta^2 + \alpha_1^2\alpha_2^2 + \alpha_1^2\delta^2 - 2\alpha_1\alpha_2\delta + 2\alpha_1^3\alpha_2\delta\right)$$

which shows that

$$b_1 = \beta_1$$

Now we can turn to $b_2$. From above, we had

$$b_2 = \frac{(r_{X_2Y} - a_2 r_{GY} - b_1 Psi'_{12})}{(1 - a_2^2)}$$

We can now directly replace $b_1$ with $\beta_1$, and we replace with the correlation terms.

$$b_2(1 - r_{X_2G}^2) = r_{X_2Y} - r_{X_2G} r_{GY} - \beta_1(r_{X_1X_2} - r_{X_1G} r_{X_2G})$$

We then replace all correlation terms with their expressions with true betas. For the left side of the equation, we have

$$b_2(1 - r_{X_2G}^2) = b_2\left(1 - (\alpha_2 + \alpha_1\delta)^2\right)$$
$$= b_2\left(1 - \alpha_2^2 - 2\alpha_2\alpha_1\delta - \alpha_1^2\delta^2\right)$$

For the right side of the equation, we have

$$
\begin{aligned}
& r_{X_2Y} - r_{X_2G} r_{GY} - \beta_1 ( r_{X_1X_2} - r_{X_1G} r_{X_2G} ) \\
&= \beta_2 + \alpha_2\gamma + \alpha_2\alpha_1\beta_1 + \delta\alpha_1\gamma + \delta\beta_1 - \left[ (\alpha_2 + \alpha_1\delta)(\gamma + \alpha_1\beta_1 + \alpha_2\beta_2 + \alpha_1\delta\beta_2) \right] - \left[ \beta_1 \left( \delta + \alpha_1\alpha_2 - (\alpha_1(\alpha_2 + \alpha_1\delta)) \right) \right] \\
&= \beta_2 + \alpha_2\gamma + {}_2\alpha_1\beta_1 + \delta\alpha_1\gamma + \delta\beta_1 - \alpha_2\gamma - \alpha_1\alpha_2\beta_1 - \alpha_2{}^2\beta_2 - \alpha_1\alpha_2\delta\beta_2 - \alpha_1\delta\gamma - \alpha_1{}^2\delta\beta_1 - \alpha_1\alpha_2\delta\beta_2 - \alpha_1{}^2\delta^2\beta_2 - \beta_1\delta + \alpha_1{}^2\delta\beta_1
\end{aligned}
$$

This can be reordered and reduced to

$$\beta_2 \left( 1 - \alpha_2{}^2 - 2\alpha_1\alpha_2\delta - \alpha_1{}^2\delta^2 \right)$$

Combining both sides of the equation, we get

$$b_2 \left( 1 - \alpha_2{}^2 - 2\alpha_2\alpha_1\delta - \alpha_1{}^2\delta^2 \right) = \beta_2 \left( 1 - \alpha_2{}^2 - 2\alpha_1\alpha_2\delta - \alpha_1{}^2\delta^2 \right),$$

which shows that

$$b_2 = \beta_2,$$

so both $b_1$ and $b_2$ are **unbiased** estimates of $\beta_1$ and $\beta_2$, respectively.

Finally, we turn to the last path $c$, which was

$$c = r_{GY} - a_1 b_1 - a_2 b_2$$

Replacing $r_{GY}$, $a_1$, $a_2$, $b_1$ and $b_2$ with the equations that we solved above, we get

$$
\begin{aligned}
c &= \gamma + \alpha_1\beta_1 + \alpha_2\beta_2 + \alpha_1\delta\beta_2 - \alpha_1\beta_1 - (\alpha_2 + \alpha_1\delta)\beta_2 \\
&= \gamma
\end{aligned}
$$

Showing that $c$ is also an **unbiased** estimate of $\gamma$ in this scenario.

Finally, define the quantities of interest (confounding, mediation effect, etc.) given the true betas. To summarise we have:

$$
\begin{aligned}
a_1 &= \alpha_1 \\
a_2 &= \alpha_2 + \alpha_1\delta \\
b_1 &= \beta_1 \\
b_2 &= \beta_2 \\
c &= \gamma
\end{aligned}
$$

with only $a_2$ being biased.

The fact that $b_1$ and $b_2$ are unbiased is very important as it means that the residual associations are unbiased even if we don't know the underlying causal relationships between exposures.

**Exposure-mediated genetic effect**

$$M_{total} = a_1 b_1 + a_2 b_2$$
$$= \alpha_1 \beta_1 + \alpha_2 \beta_2 + \alpha_1 \delta \beta_2$$

This corresponds to the effect mediated in the true causal model including via the pathway: $G \rightarrow X_1 \rightarrow X_2 \rightarrow Y$. So, the total mediation effect is **unbiased**. This is an important result as the constraint we impose for the definition of the heritability is equal to

$$M_{total} + c = \alpha_1 \beta_1 + \alpha_2 \beta_2 + \alpha_1 \delta \beta_2 + \gamma$$

which also corresponds to all pathways from *G* to *Y* in the true model.

The effect mediated by a specific pathway however can be biased:
Mediation via $X_1$ is **unbiased**, as we have

$$a_1 b_1 = \alpha_1 \beta_1$$

However, mediation via $X_2$ will be **biased** if the causal effect $\delta$ is not specified:

$$a_2 b_2 = \alpha_2 \beta_2 + \alpha_1 \delta \beta_2$$

If we don't know the causal structure between the exposures, we also don't know which mediation effect is biased. That said, the bias $\alpha_1\delta\beta_2$ is still mediated via $X_2$, but it is possible that the fitted mediation effect is significant even if $\alpha_2$ = 0. To illustrate this bias from an intervention perspective, let's assume $\alpha_2$ = 0. In such case, it would be theoretically sufficient to intervene on $X_1$, which would block both indirect paths $\alpha_1\beta_1$ and $\alpha_1\delta\beta_2$. However, our fitted model here would suggest intervening both on $X_1$ and $X_2$.

**Genetic confounding**

For the $X_1$-$Y$ association, this is

$$C_1 = a_1 a_2 b_2 + a_1 c$$
$$= \alpha_1 (\alpha_2 + \alpha_1 \delta) \beta_2 + \alpha_1 \gamma$$

So

$$C_1 = a_1 a_2 b_2 + a_1 c = \alpha_1 \alpha_2 \beta_2 + \alpha_1 \gamma + \alpha_1^2 \delta \beta_2 ,$$

Where the estimated $a_2$ path is a combination of the true $\alpha_1$ path and the mediation via $X_1$ ($\alpha_1\delta$), this becomes part of the genetic confounding.

For the $X_2$-$Y$ association, this is

$$C_2 = a_2\gamma + \alpha_2\alpha_1\beta_1 + \delta\alpha_1\gamma + \delta\alpha_1{}^2\beta_1$$

**Genetic overlap**

For the $X_1$-$Y$ association, this is

$$O_1 = C_1 + a_1{}^2 b_1 = C_1 + \alpha_1{}^2\beta_1$$

As the confounding paths already contain the path including $\delta$, this is not further specified here.

For the $X_2$-$Y$ association, this is

$$\begin{aligned} O_2 &= C_2 + a_2{}^2 b_2 \\ &= C_2 + (\alpha_2{}^2 + 2\alpha_2\alpha_1\delta + \alpha_1{}^2\delta^2)\beta_2 \end{aligned}$$

Let's remember that in the true model, the variance of $X_1$ is

$$V_{X_1} = \alpha_1{}^2 V_G + V_{\varepsilon_{X_1}}$$

and

$$V_{X_2} = \alpha_2{}^2 V_G + 2\alpha_2\alpha_1\delta V_G + \delta^2\alpha_1{}^2 V_G + \delta^2 V_{\varepsilon_{X_1}} + V_{\varepsilon_{X_2}}$$

So, we have

$$\alpha_2{}^2 V_G + 2\alpha_2\alpha_1\delta V_G + \delta^2\alpha_1{}^2 V_G = V_{X_2} - \delta^2 V_{\varepsilon_{X_1}} - V_{\varepsilon_{X_2}}$$

The left expression is therefore all the components of the variance in $X_2$ that originate in *G*. And the right expression is all the non-genetic residual variance in $X_2$.

So, the expression

$$(\alpha_2{}^2 + 2\alpha_2\alpha_1\delta + \alpha_1{}^2\delta^2)\beta_2$$

corresponds indeed to the part to the causal effect scaled by the variance originating in *G*, which corresponds to the causal part of the genetic overlap.

#### 4. Unobserved environmental confounder in the true model

In this section, we show how the fitted *Gsens* model (accounting for measurement error in *g*) can be used to accurately estimate the effects of the true model. For simplicity, this section assumes no measurement error in *Y* and in the exposures.

**Figure S4a** The true model

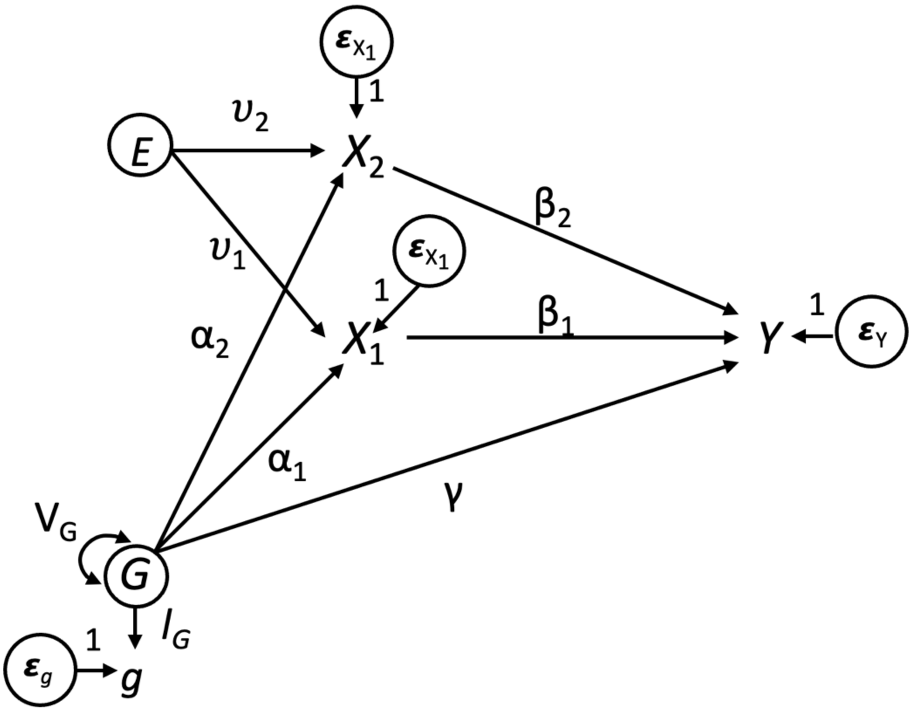


**True model equations:**

$$Y = \gamma G + \beta_1 X_1 + \beta_2 X_2 + \varepsilon_Y$$

$$X_1 = \alpha_1 G + \upsilon_1 E + \varepsilon_{X_1}$$

$$X_2 = \alpha_2 G + \upsilon_2 E + \varepsilon_{X_2}$$

$$g = l_G G + \varepsilon_g$$

$$G = G$$

#### Additional assumptions

- When there is a common cause *E* with effects on $X_1$ and $X_2$, error terms $e_{X1}$ and $e_{X2}$ are expected to be correlated in the fitted model
- This section assumes that $l_G$ in the fitted model equals $l_G$ in the true model, which in reality cannot be verified

**Figure S4b** The fitted model

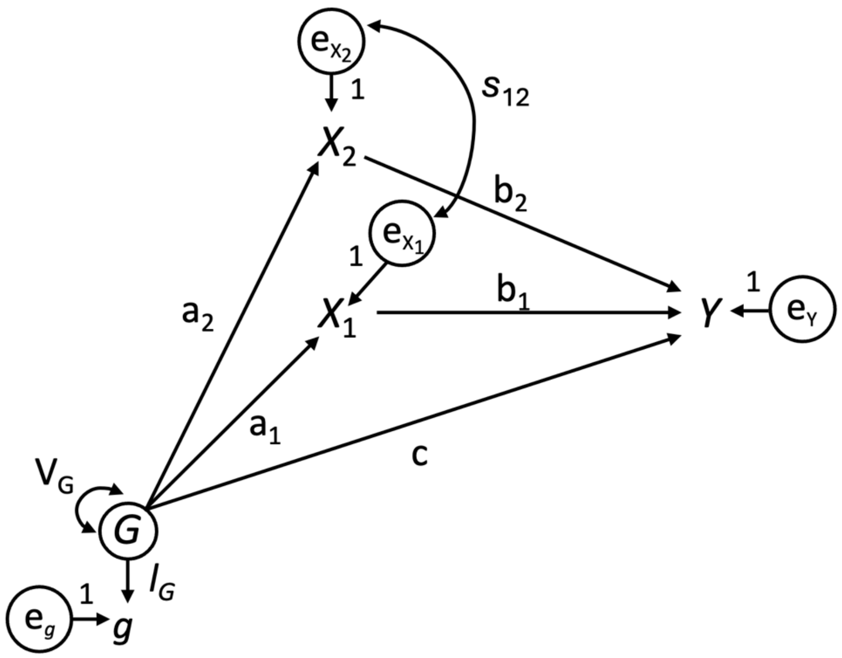


**Fitted model equations –** assuming no measurement error in *Y* and *X*s, but allowing residual correlation between exposures

$$Y = cG + b_1X_1 + b_2X_2 + e_Y$$

$$X_1 = a_1G + e_{X_1}$$

$$X_2 = a_2G + e_{X_2}$$

$$g = l_GG + e_g$$

$$G = G$$

**Derive the variances in the fitted model**

$$Var_Y = c^2V_G + 2cb_1Cov(G, X_1) + 2cb_2Cov(G, X_2) + 2b_1b_2Cov(X_1, X_2) + b_1^2V_{X_1} + b_2^2V_{X_2} + V_{e_Y}$$

$$Var_{X_1} = a_1^2V_G + V_{e_{X_1}}$$

$$Var_{X_2} = a_2^2V_G + V_{e_{X_2}}$$

**Derive the covariances in the fitted model**

For the covariances between exposures and the *G* we have

$$Cov(X_1, G) = Cov(a_1G + e_{X_1}, G) = a_1V_G$$

$$Cov(X_2, G) = Cov(a_2G + e_{X_2}, G) = a_2V_G$$

which when standardised correspond to:

$$r_{X_1G} = a_1$$

$$r_{X_2G} = a_2$$

The covariance between exposure 1 ($X_1$) and the outcome $Y$ is

$$Cov(X_1, Y) = a_1cV_G + a_1b_1Cov(G, X_1) + a_1b_2Cov(G, X_2) + b_1Cov(e_{X_1}, X_1) + b_2Cov(e_{X_1}, X_2)$$

In general, we assume that the error term of one variable is unrelated to another variable and that error terms of two variables are unrelated (e.g., error terms of $G$ and $Y$). However, as we do not model any environmental confounder between exposures, we assume the error terms of $X_1$ and $X_2$ to be correlated, i.e.,

$$Cov(e_{X_1}, e_{X_2}) \neq 0$$

Now we replace $X_1$ and $X_2$ to simplify the covariance between $X_1$ and $Y$

$$Cov(X_1, Y) = b_1(a_1^2V_G + V_{e_{X_1}}) + a_1cV_G + a_1a_2b_2V_G + b_2Cov(e_{X_1}, e_{X_2})$$

where

$$V_{e_{X_1}} + a_1^2V_G = V_{X_1}$$

Thus, we have

$$Cov(X_1, Y) = b_1V_{X_1} + a_1cV_G + a_1a_2b_2V_G + b_2Cov(e_{X_1}, e_{X_2}) \quad .$$

Moving forward, the residual covariance between $X_1$ and $X_2$ in the fitted model will be referred to as $s_{12}$ and the residual correlation as $s'_{12}$. When standardised, the correlation between $X_1$ and $Y$ is:

$$r_{X_1Y} = b_1 + a_1c + a_1a_2b_2 + b_2s'_{12}.$$

Similarly, for the covariance between $X_2$ and $Y$, we have:

$$Cov(X_2, Y) = b_2V_{X_2} + a_2cV_G + a_2a_1b_1V_G + b_1Cov(e_{X_1}, e_{X_2})$$

and

$$r_{X_2Y} = b_2 + a_2c + a_2a_1b_1 + b_1s'_{12}$$

For the covariance and correlation between exposures 1 and 2 we have

$$\begin{aligned} Cov\left(X_1, X_2\right) &= Cov\left(a_1G + e_{X_1}, a_2G + e_{X_2}\right) \\ &= a_1a_2V_G + s_{12} \end{aligned}$$

and

$$\begin{aligned} r_{X_1X_2} &= a_1a_2 + s'_{12} \\ &= r_{X_1G}r_{X_2G} + s'_{12} \end{aligned}$$

From the equations above we get the following equations:

$$s'_{12} = r_{X_1X_2} - r_{X_1G}r_{X_2G} \quad (4.1)$$

$$b_1 = r_{X_1Y} - a_1c - a_1a_2b_2 - b_2s'_{12} \quad (4.2)$$

$$b_2 = r_{X_2Y} - a_2c - a_2a_1b_1 - b_1s'_{12} \quad (4.3)$$

$$c = r_{GY} - a_1b_1 - a_2b_2 \quad (4.4)$$

**Step 2. Express betas (fitted model) as a function of observed correlations.**

Now we replace *c* in equation (4.2) with equation (4.4), which is

$$\begin{aligned} b_1 &= r_{X_1Y} - a_1\left(r_{GY} - a_1b_1 - a_2b_2\right) - a_1a_2b_2 - b_2s'_{12} \\ &= r_{X_1Y} - a_1r_{GY} + a_1^{\,2}b_1 - b_2s'_{12} \end{aligned}$$

$$b_1\left(1 - a_1^{\,2}\right) = r_{X_1Y} - a_1r_{GY} - b_2s'_{12} \quad (4.5a)$$

$$b_1 = \frac{r_{X_1Y} - a_1r_{GY} - b_2s'_{12}}{\left(1 - a_1^{\,2}\right)} \quad (4.5b)$$

Replacing *c* from equation (4.4) can also be done for equation (4.3), which equivalently leads to

$$b_2 = \frac{r_{X_2Y} - a_2r_{GY} - b_1s'_{12}}{\left(1 - a_2^{\,2}\right)} \quad (4.6)$$

Next, we can replace $b_2$ in equation (4.5a) with the expression we derived from equation (4.6), which is

$$b_1\left(1 - a_1^2\right) = r_{X_1Y} - a_1 r_{GY} - s'_{12}\frac{r_{X_2Y} - a_2 r_{GY} - b_1 s'_{12}}{\left(1 - a_2^2\right)}$$

We now replace $a_1$, $a_2$ and $s'_{12}$ by their respective equations from above, we arrive at

$$b_1\left(1 - r_{X_1G}^2 - r_{X_2G}^2 - r_{X_1X_2}^2 + 2r_{X_1X_2}r_{X_1G}r_{X_2G}\right) = r_{X_1Y} - r_{X_1Y}r_{X_2G}^2 - r_{X_1G}r_{GY} - r_{X_1X_2}r_{X_2Y} + r_{X_1G}r_{X_2G}r_{X_2Y} + r_{X_1X_2}r_{X_2G}r_{GY}$$

Now we can turn to the true model (**Figure S4a**).

**Define variances in the true model**

$$\begin{aligned} Var\left(X_1\right) &= Cov\left(\alpha_1 G + \upsilon_1 E + \varepsilon_{X_1}, \alpha_1 G + \upsilon_1 E + \varepsilon_{X_1}\right) \\ &= \alpha_1^2 V_G + \upsilon_1^2 V_E + V_{\varepsilon_{X_1}} \end{aligned}$$

$$Var\left(X_2\right) = \alpha_2^2 V_G + \upsilon_2^2 V_E + V_{\varepsilon_{X_2}}$$

$$Var(Y) = \gamma^2 V_G + 2\gamma\beta_1\alpha_1 V_G + 2\gamma\beta_2\alpha_2 V_G + 2\beta_1\beta_2\alpha_1\alpha_2 V_G + \beta_1^2 V_{X_1} + \beta_2^2 V_{X_2} + V_{\varepsilon_Y}$$

**Covariances in the true model**

$$Cov\left(X_1, G\right) = \alpha_1 V_G$$

$$Cov\left(X_2, G\right) = \alpha_2 V_G$$

$$Cov\left(X_1, X_2\right) = \alpha_1\alpha_2 V_G + \upsilon_1\upsilon_2 V_E$$

$$Cov\left(X_1, Y\right) = \alpha_1\gamma V_G + \beta_1 V_{X_1} + \beta_2\left(\alpha_1\alpha_2 V_G + \upsilon_1\upsilon_2 V_E\right)$$

which when standardised is

$$r_{X_1Y} = \alpha_1\gamma + \beta_1 + \beta_2\left(\alpha_1\alpha_2 + \upsilon_1\upsilon_2\right)$$

Similarly, the covariance and correlation between $X_2$ and $Y$ are

$$Cov\left(X_2, Y\right) = \alpha_2\gamma V_G + \beta_2 V_{X_2} + \beta_1\left(\alpha_1\alpha_2 V_G + \upsilon_1\upsilon_2 V_E\right)$$

and

$$r_{X_2Y} = \alpha_2\gamma + \beta_2 + \beta_1\left(\alpha_1\alpha_2 + \upsilon_1\upsilon_2\right)$$

*Covariance between G and Y*

$$Cov(G,\ Y) = \gamma V_G + \alpha_1\beta_1 V_G + \alpha_2\beta_2 V_G$$

and standardised

$$r_{GY} = \gamma + \alpha_1\beta_1 + \alpha_2\beta_2$$

So, from this subsection we have the following equations:

$$r_{X_1G} = \alpha_1$$
$$r_{X_2G} = \alpha_2$$
$$r_{X_1X_2} = \alpha_1\alpha_2 + \upsilon_1\upsilon_2$$
$$r_{X_1Y} = \alpha_1\gamma + \beta_1 + \beta_2\left(\alpha_1\alpha_2 + \upsilon_1\upsilon_2\right)$$
$$r_{X_2Y} = \alpha_2\gamma + \beta_2 + \beta_1\left(\alpha_1\alpha_2 + \upsilon_1\upsilon_2\right)$$
$$r_{GY} = \gamma + \alpha_1\beta_1 + \alpha_2\beta_2$$

**Step 4. Express betas (fitted model) as a function of the true betas.**

For $b_1$, we had the following equation

$$b_1\left(1 - r_{X_1G}{}^2 - r_{X_2G}{}^2 - r_{X_1X_2}{}^2 + 2r_{X_1X_2}r_{X_1G}r_{X_2G}\right) = r_{X_1Y} - r_{X_1Y}r_{X_2G}{}^2 - r_{X_1G}r_{GY} - r_{X_1X_2}r_{X_2Y} + r_{X_1G}r_{X_2G}r_{X_2Y} + r_{X_1X_2}$$ [illegible]

We begin with solving the left part of the equation, which is

$$\begin{aligned} & b_1\left(1 - r_{X_1G}{}^2 - r_{X_2G}{}^2 - r_{X_1X_2}{}^2 + 2r_{X_1X_2}r_{X_1G}r_{X_2G}\right) \\ &= b_1\left[1 - \alpha_1{}^2 - \alpha_2{}^2 - \left(\alpha_1\alpha_2 + \upsilon_1\upsilon_2\right)^2 + 2\left(\alpha_1\alpha_2 + \upsilon_1\upsilon_2\right)\alpha_1\alpha_2\right] \\ &= b_1\left(1 - \alpha_1{}^2 - \alpha_2{}^2 - \alpha_1{}^2\alpha_2{}^2 - \upsilon_1{}^2\upsilon_2{}^2 - 2\alpha_1\alpha_2\upsilon_1\upsilon_2 + 2\alpha_1{}^2\alpha_2{}^2 + 2\upsilon_1\upsilon_2\alpha_1\alpha_2\right) \\ &= b_1\left(1 - \alpha_1{}^2 - \alpha_2{}^2 - \upsilon_1{}^2\upsilon_2{}^2 + \alpha_1{}^2\alpha_2{}^2\right) \end{aligned}$$

For the right side of the equation, we have

$$\begin{aligned} & r_{X_1Y} - r_{X_1Y}r_{X_2G}{}^2 - r_{X_1G}r_{GY} - r_{X_1X_2}r_{X_2Y} + r_{X_1G}r_{X_2G}r_{X_2Y} + r_{X_1X_2}r_{X_2G}r_{GY} \\ &= \beta_1\left(1 - \alpha_1{}^2 - \alpha_2{}^2 - \upsilon_1{}^2\upsilon_2{}^2 + \alpha_1{}^2\alpha_2{}^2\right) \end{aligned}$$

Bringing together both sides of the equation we get

$$b_1\left(1-\alpha_1^2-\alpha_2^2-\upsilon_1^2\upsilon_2^2+\alpha_1^2\alpha_2^2\right)=\beta_1\left(1-\alpha_1^2-\alpha_2^2-\upsilon_1^2\upsilon_2^2+\alpha_1^2\alpha_2^2\right),$$

Showing that

$$b_1=\beta_1$$

Equivalently, we have

$$b_2\left(1-\alpha_1^2-\alpha_2^2-\upsilon_1^2\upsilon_2^2+\alpha_1^2\alpha_2^2\right)=\beta_2\left(1-\alpha_1^2-\alpha_2^2-\upsilon_1^2\upsilon_2^2+\alpha_1^2\alpha_2^2\right),$$

So

$$b_2=\beta_2$$

That means that both $b_1$ and $b_2$ are **unbiased** estimates of $\beta_1$ and $\beta_2$, respectively. As $a_1$ and $a_2$ are also **unbiased** estimates of $\alpha_1$ and $\alpha_2$ in this scenario, the total mediation and individual mediation paths will also be **unbiased**.

Finally, to show that genetic confounding and genetic overlap are unbiased, we can show that $c$ is also an unbiased estimate of $\gamma$ as from equation 3.4 we had

$$c=r_{GY}-a_1b_1-a_2b_2$$

We can now insert the equation for $r_{GY}$ and replace $a_1$, $a_2$ $b_1$ and $b_2$ with the true parameters and we get

$$c=\gamma+\alpha_1\beta_1+\alpha_2\beta_2-\alpha_1\beta_1-\alpha_2\beta_2$$

$$c=\gamma$$

### 5. Equivalence of equality constraints – heritability vs ratio

In the first paper where measurement error in polygenic scores was conceptualised, one approach was to use structural equation models with a latent genetic factor (Tucker-Drob, 2017). The author proposed the following ratio to capture and correct for measurement error:

$$l_G{}^2 = \frac{r_{GY}{}^2}{h^2{}_{SNP}} \qquad (5.1)$$

Similar specifications were used in other papers (Becker et al., 2021). In our review article (Pingault et al., 2022), we showed how this ratio made assumptions in particular regarding measurement error in the outcome.

In the initial *Gsens* paper (Pingault et al., 2021), we corrected for measurement error in our one-exposure model by imposing an equality constraint, which in the present notation is

$$\gamma + \alpha_1\beta_1 = \sqrt{h^2{}_{SNP}} \qquad (5.2),$$

As we show below, these constraints are equivalent. From equation 5.1 we get

$$\sqrt{h^2{}_{SNP}}\, l_G = r_{GY} \qquad (5.3)$$

In the one-exposure case, we get from previous derivations and path tracing rules:

$$r_{gY} = l_G(\gamma + \alpha_1\beta_1) \qquad (5.4)$$

Replacing $r_{GY}$ in equation 5.3 with $r_{gY}$ from equation 5.4, we get

$$\sqrt{h^2{}_{SNP}}\, l_G = r_{gY} = l_G\left(\gamma + \alpha_1\beta_1\right)$$

simplifying to

$$\sqrt{h^2{}_{SNP}} = \gamma + \alpha_1\beta_1,$$

showing that the loading constraint (5.1) and the heritability constraint (5.2) are equivalent.

This equivalence generalises to whatever paths from *G* to *Y* constitute heritability, including via multiple exposures.

If we note $\sum_{i=1}^{p} P_i$ the sum of all legitimate directed paths from *G* to *Y*. To reach *g*, each of those path $P_i$ must be multiplied by $l_G$, which can thus be factored out of the sum, so we get:

$$r_{GY} = 1_G \sum_{i=1}^{p} P_i$$

So, starting again from the ratio constraint as expressed in 5.3:

$$\sqrt{h_{SNP}}\, 1_G = r_{GY}$$

We find, after simplifying by $l_G$ on both sides:

$$\sqrt{h_{SNP}} = \sum_{i=1}^{p} P_i$$

which is the equivalent of our heritability constraint, as under our assumption that $G$ = $G_{SNP}$ the SNP heritability comprises the sum of all legitimate paths from $G$ to $Y$.

Whatever the form that the heritability of $Y$ takes, the ratio and the heritability constraint are thus equivalent (i.e. whether there is only one direct path $\gamma$ from $G$ to $Y$ as the original model from Tucker-Drob, 2017, or whether heritability is also mediated one trait or several traits).

**6. Bias amplification (collider bias)**

As opposed to using the 'true' genetic factor for the outcome, which is what we do in *Gsens*, one may consider using the true genetic factor for *X* to account for genetic confounding. However, the true genetic factor for *X* will include genetic variants that do not have any effects on *Y*, i.e., these are genetic instruments for *X* and associations between these instruments and *Y* will only occur through *X*. Here we illustrate in which scenarios this approach becomes problematic. Let us assume the following true and fitted models:

**Figure S6a** The true model

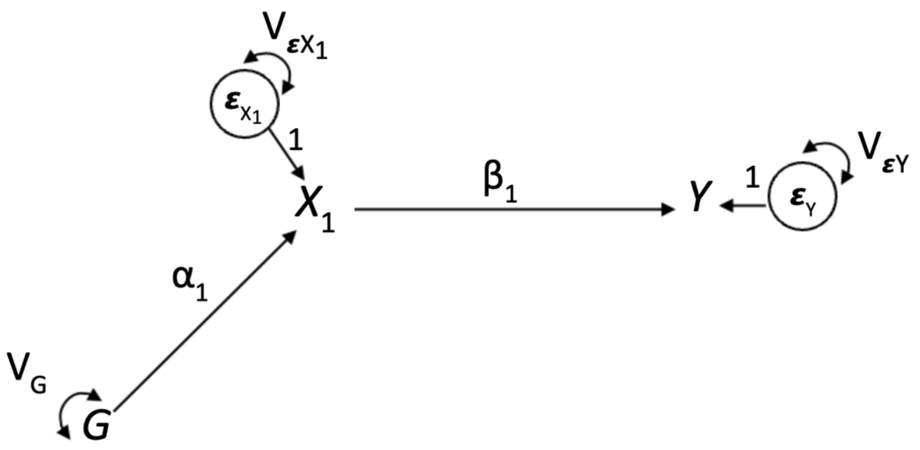


**Figure S6b** The fitted model

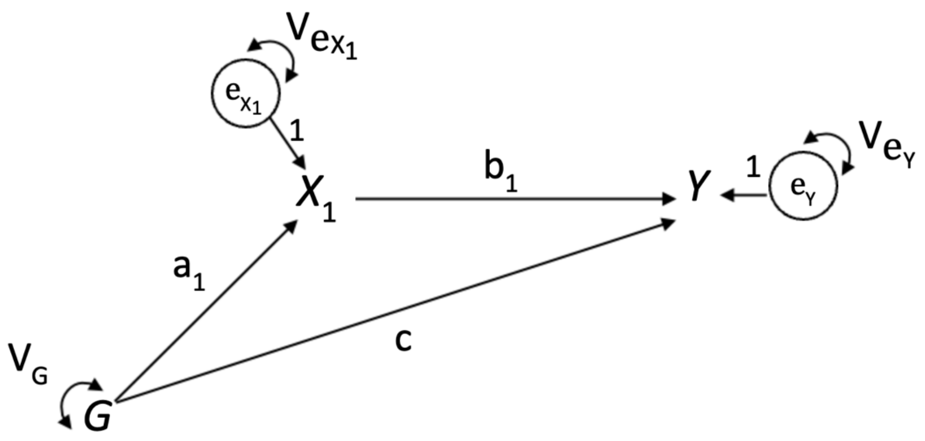


Using a similar approach to find fitted parameters as a function of true parameters, we can show, that, as expected: $b_1 = \beta_1$

Therefore, we get back the true beta even if we fit a model that adjust for *G* when *G* is not a confounder. So, there **no bias amplification**. Now let us add an additional variable *U*, but again no confounding in the true model, which results in $X_1$ being a **collider**, which becomes the following **true model.**

**Figure S6c** True model with collider

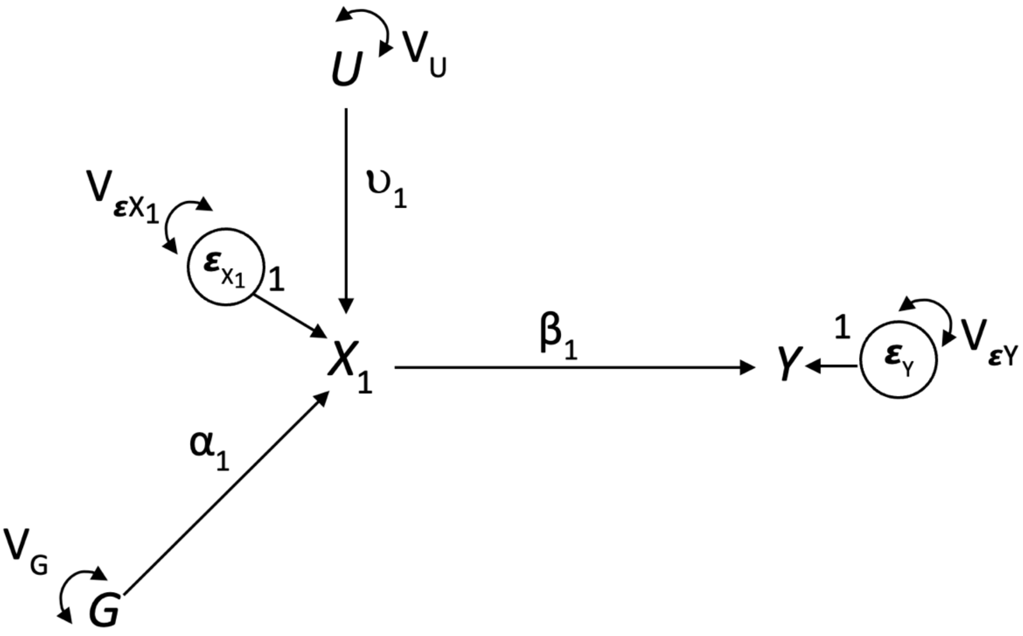


Here, we can show again that:

$$b_1 = \beta_1$$

So, the addition of a collider per se, in the sense that $U$ and $G$ collide in $X_1$ and that we adjust for $X_1$ in the fitted model does not lead to bias. Now let us add the confounding path from $U$:

**Figure S6c** True model with collider and confounding

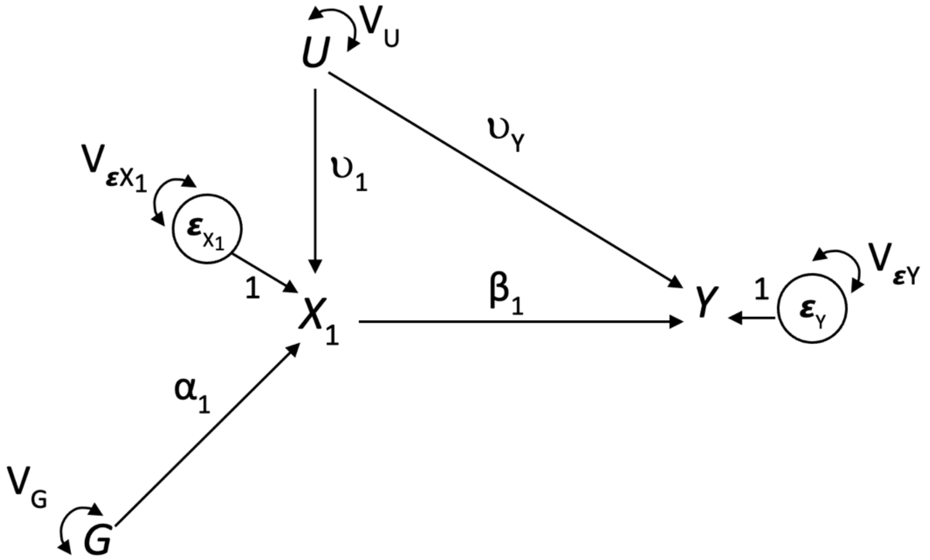

In this case, as there is no confounding from *G*, the true residual association after accounting for *G* should be:

$$b_1 = r_{X_1Y} = \beta_1 + \upsilon_1\upsilon_Y$$

In other words, we should retrieve the correlation between $X_1$ and *Y*, which should be unaffected by the adjustment for a non-confounder.

Instead, we can show that

$$b_1 = \beta_1 + \frac{\upsilon_1\upsilon_Y}{(1 - \alpha_1^2)}$$

Meaning that the bias coming from the unobserved confounder is amplified by the denominator. Bias amplification thus occurs when adjusting for a variable that behaves as an instrument of the exposure in the presence of unknown/unadjusted confounding of the exposure outcome relationship. When *G* is a typical instrument for $X_1$ (e.g. a SNP), bias amplification is not too serious, as typically very little variance in $X_1$ is explained by *G* (i.e., $\alpha_1^2$ will be very small) (Myers et al., 2011). However, it does matter for *Gsens* where the polygenic score, and even more the polygenic score corrected for measurement error, explains a substantial part of the variance in $X_1$, leading to serious bias amplification.

### 7. Supplementary Results for the Empirical Application of Gsens in MoBa

#### Supplementary Tables

**Supplementary Table S1**

*Descriptive Statistics of the MoBa Sample*

| | **Full sample** | | | | **Genotyped sample** | | | |
|---|---|---|---|---|---|---|---|---|
| **Variable** | *N* | Mean | *SD* | Range | *N* | Mean | *SD* | Range |
| ADHD 8y | 43,224 | 8.54 | 7.23 | 0–54 | 31,476 | 8.40 | 7.10 | 0–54 |
| Birthweight (in g) | 113,117 | 3553.5 | 616.4 | 100–6320 | 76,207 | 3615.7 | 544.7 | 260–6320 |
| Temperament (Emotionality) | 58,149 | 5.38 | 2.31 | 0–12 | 42,644 | 5.37 | 2.30 | 0–12 |
| Temperament (Activity) | 58,192 | 7.87 | 2.11 | 0–12 | 42,674 | 7.87 | 2.10 | 0–12 |
| Maternal SDP (start of pregnancy) | 95,002 | 1.23 | 0.62 | 1–3 | 64,408 | 1.22 | 0.61 | 1–3 |
| Maternal SDP (after pregnancy known) | 91,705 | 1.13 | 0.48 | 1–3 | 62,316 | 1.12 | 0.46 | 1–3 |
| Maternal BMI (pre-pregnancy) | 98,851 | 23.83 | 3.91 | 17–45 | 69,530 | 23.85 | 3.87 | 17–44 |
| Maternal education | 97,708 | 5.56 | 1.27 | 2–7 | 68,720 | 5.59 | 1.24 | 2–7 |
| Paternal education | 93,611 | 5.21 | 1.45 | 2–7 | 66,049 | 5.23 | 1.44 | 2–7 |

**Supplementary Table S2**

*Phenotypic Associations Between Predisposing Factors and ADHD*

| **Model 1** | | | | |
|---|---|---|---|---|
| Individual factor | *Std Est* | *SE* | *t* | *p*-value |
| Sex | –0.18 | 0.006 | –29.50 | $4.0 \times 10^{-188}$ |
| Birth weight | –0.05 | 0.006 | –7.59 | $3.3 \times 10^{-14}$ |
| EAS Emotionality | 0.18 | 0.006 | 30.83 | $4.5 \times 10^{-205}$ |
| EAS Activity | 0.14 | 0.006 | 24.18 | $8.4 \times 10^{-128}$ |

| **Model 2** | | | | |
|---|---|---|---|---|
| Parental exposure | *Std Est* | *SE* | *t* | *p*-value |
| Sex | –0.17 | 0.007 | –25.74 | $6.6 \times 10^{-144}$ |
| Maternal education | –0.03 | 0.008 | –3.94 | $8.0 \times 10^{-05}$ |
| Paternal education | –0.05 | 0.008 | –5.77 | $8.0 \times 10^{-09}$ |
| SDP (beginning) | 0.04 | 0.009 | 5.03 | $5.0 \times 10^{-07}$ |
| Smoking during pregn. | 0.02 | 0.009 | 2.39 | 0.017 |
| Maternal BMI (pregn.) | 0.03 | 0.007 | 4.03 | $5.7 \times 10^{-05}$ |

*Note. Std Est* is the standardised regression coefficient. For model 2, effects of the covariates maternal and paternal age are not shown to align with the MoBa policy of not publishing effects of confounders.

**Supplementary Table S3**

*Gsens Results for Example 1 – Birth Weight, Emotionality and Activity* ***(Unrelated Individuals)***

| | *Sdt Est* | *SE* | CI lower | CI upper | *Z* | *p*-value |
|---|---|---|---|---|---|---|
| Sex | –0.191 | 0.012 | –0.213 | –0.168 | –16.28 | $1.3 \times 10^{-59}$ |
| Birth weight | –0.032 | 0.011 | –0.054 | –0.010 | –2.80 | $5.1 \times 10^{-03}$ |
| EAS Emotionality | 0.172 | 0.011 | 0.150 | 0.194 | 15.55 | $1.5 \times 10^{-54}$ |
| EAS Activity | 0.065 | 0.013 | 0.040 | 0.090 | 5.07 | $4.0 \times 10^{-07}$ |
| Mediation via BW | 0.001 | 0.001 | 0.000 | 0.002 | 2.15 | 0.031 |
| Mediation via EAS_Emotionality | 0.009 | 0.004 | 0.001 | 0.017 | 2.13 | 0.033 |
| Mediation via EAS_Activity | 0.015 | 0.002 | 0.011 | 0.018 | 7.81 | $5.5 \times 10^{-15}$ |
| Total mediation | 0.021 | 0.007 | 0.006 | 0.035 | 2.86 | $4.3 \times 10^{-03}$ |
| Genetic confounding BW | –0.015 | 0.010 | –0.034 | 0.005 | –1.49 | 0.137 |
| Genetic confounding EAS_Emo | 0.018 | 0.009 | 0.000 | 0.037 | 2.00 | 0.048 |
| Genetic confounding EAS_Act | 0.082 | 0.010 | 0.061 | 0.102 | 7.86 | $3.7 \times 10^{-15}$ |
| Genetic overlap BW | –0.191 | 0.012 | –0.213 | –0.168 | –16.28 | $1.3 \times 10^{-59}$ |
| Genetic overlap EAS_Emo | –0.032 | 0.011 | –0.054 | –0.010 | –2.80 | $5.1 \times 10^{-03}$ |
| Genetic overlap EAS_Act | 0.172 | 0.011 | 0.150 | 0.194 | 15.55 | $1.5 \times 10^{-54}$ |

*Note.* 95% confidence intervals are specified. BW = birth weight, EAS = Emotionality, Activity and Shyness Scale

**Supplementary Table S4**

*Gsens Results for Example 1 – Birth Weight, Emotionality and Activity* ***(Causal Effect of Birth Weight on Emotionality)***

| | *Sdt Est* | *SE* | CI lower | CI upper | *Z* | *p*-value |
|---|---|---|---|---|---|---|
| Sex | –0.190 | 0.009 | –0.208 | –0.172 | –20.44 | $7.6 \times 10^{-93}$ |
| Birth weight | –0.033 | 0.009 | –0.050 | –0.015 | –3.63 | $2.8 \times 10^{-04}$ |
| EAS Emotionality | 0.170 | 0.009 | 0.153 | 0.187 | 19.42 | $5.7 \times 10^{-84}$ |
| EAS Activity | 0.062 | 0.010 | 0.042 | 0.082 | 6.12 | $9.3 \times 10^{-10}$ |
| Mediation via BW | 0.001 | 0.000 | 0.001 | 0.002 | 3.10 | $1.9 \times 10^{-03}$ |
| Mediation via EAS_Emotionality | 0.009 | 0.003 | 0.003 | 0.015 | 2.83 | $5.8 \times 10^{-03}$ |
| Mediation via EAS_Activity | 0.014 | 0.002 | 0.011 | 0.017 | 9.22 | $2.9 \times 10^{-20}$ |
| **BW → EAS_Emotionality** | **–0.024** | 0.006 | –0.037 | –0.012 | –3.88 | $1.1 \times 10^{-04}$ |
| **Mediation BW → EAS_Emo** | **–0.004** | 0.001 | –0.006 | –0.002 | –3.73 | $1.9 \times 10^{-04}$ |
| **Total mediation** | **0.019** | 0.006 | 0.008 | 0.031 | 3.41 | $6.6 \times 10^{-04}$ |
| Genetic confounding BW | –0.016 | 0.008 | –0.032 | –0.001 | –2.12 | 0.034 |
| Genetic confounding EAS_Emo | 0.019 | 0.007 | 0.005 | 0.033 | 2.62 | $8.8 \times 10^{-03}$ |
| Genetic confounding EAS_Act | 0.082 | 0.008 | 0.066 | 0.098 | 10.02 | $1.2 \times 10^{-23}$ |
| Genetic overlap BW | –0.190 | 0.009 | –0.208 | –0.172 | –20.44 | $7.6 \times 10^{-93}$ |
| Genetic overlap EAS_Emo | –0.033 | 0.009 | –0.050 | –0.015 | –3.63 | $2.8 \times 10^{-04}$ |
| Genetic overlap EAS_Act | 0.170 | 0.009 | 0.153 | 0.187 | 19.42 | $5.7 \times 10^{-84}$ |

**Supplementary Table S5**

*Gsens Results for Example 1 – Birth Weight, Emotionality and Activity* ***(Causal Effect of Birth Weight on Activity)***

| | *Std Est* | *SE* | CI lower | CI upper | *Z* | *p*-value |
|---|---|---|---|---|---|---|
| Sex | –0.190 | 0.009 | –0.208 | –0.172 | –20.44 | $7.6 \times 10^{-93}$ |
| Birth weight | –0.033 | 0.009 | –0.050 | –0.015 | –3.63 | $2.8 \times 10^{-04}$ |
| EAS Emotionality | 0.170 | 0.009 | 0.153 | 0.187 | 19.42 | $5.7 \times 10^{-84}$ |
| EAS Activity | 0.062 | 0.010 | 0.042 | 0.082 | 6.12 | $9.3 \times 10^{-10}$ |
| Mediation via BW | 0.001 | 0.000 | 0.001 | 0.002 | 3.10 | $1.9 \times 10^{-03}$ |
| Mediation via EAS_Emotionality | 0.009 | 0.003 | 0.003 | 0.015 | 2.83 | $4.7 \times 10^{-03}$ |
| Mediation via EAS_Activity | 0.014 | 0.002 | 0.011 | 0.017 | 9.22 | $2.7 \times 10^{-20}$ |
| **BW → EAS_Activity** | **–0.006** | 0.008 | –0.021 | 0.009 | –0.78 | 0.438 |
| **Mediation BW → EAS_Activity** | **0.000** | 0.000 | –0.001 | 0.001 | –0.76 | 0.448 |
| **Total mediation** | **0.019** | 0.006 | 0.008 | 0.031 | 3.41 | $6.6 \times 10^{-04}$ |
| Genetic confounding BW | –0.016 | 0.008 | –0.032 | –0.001 | –2.12 | 0.034 |
| Genetic confounding EAS_Emo | 0.019 | 0.007 | 0.005 | 0.033 | 2.62 | $8.8 \times 10^{-03}$ |
| Genetic confounding EAS_Act | 0.082 | 0.008 | 0.066 | 0.098 | 10.02 | $1.2 \times 10^{-23}$ |
| Genetic overlap BW | –0.190 | 0.009 | –0.208 | –0.172 | –20.44 | $7.6 \times 10^{-93}$ |
| Genetic overlap EAS_Emo | –0.033 | 0.009 | –0.050 | –0.015 | –3.63 | $2.8 \times 10^{-04}$ |
| Genetic overlap EAS_Act | 0.170 | 0.009 | 0.153 | 0.187 | 19.42 | $5.7 \times 10^{-84}$ |

**Supplementary Table S6**

*Gsens Results for Example 2 – Parental Education, Maternal Smoking and BMI During Pregnancy* ***(Unrelated Individuals)***

| | Std Est | *SE* | CI lower | CI upper | Z | *p*-value |
|---|---|---|---|---|---|---|
| Maternal education | 0.018 | 0.016 | –0.013 | 0.049 | 1.12 | 0.261 |
| Paternal education | 0.045 | 0.018 | 0.010 | 0.081 | 2.49 | 0.013 |
| Smoking during pregnancy (start) | 0.023 | 0.017 | –0.011 | 0.058 | 1.33 | 0.185 |
| Smoking during pregnancy | –0.003 | 0.018 | –0.037 | 0.032 | –0.15 | 0.878 |
| Maternal BMI (pre-pregnancy) | –0.016 | 0.014 | –0.044 | 0.012 | –1.10 | 0.270 |
| Genetic confounding Mat. Edu | –0.102 | 0.014 | –0.129 | –0.075 | –7.41 | $1.3 \times 10^{-13}$ |
| Genetic confounding Pat. Edu | –0.127 | 0.016 | –0.159 | –0.950 | –7.87 | $3.4 \times 10^{-15}$ |
| Genetic confounding SDP (start) | 0.055 | 0.011 | 0.033 | 0.077 | 4.93 | $8.1 \times 10^{-07}$ |
| Genetic confounding SDP | 0.054 | 0.011 | 0.032 | 0.077 | 4.80 | $1.6 \times 10^{-06}$ |
| Genetic confounding Mat. BMI | 0.066 | 0.012 | 0.043 | 0.090 | 5.53 | $3.2 \times 10^{-08}$ |

*Note.* This table omits the genetic overlap estimates, which are highly similar to the respective genetic confounding estimates.
